# Combining individual and wastewater whole genome sequencing improves SARS-CoV-2 surveillance

**DOI:** 10.1101/2024.07.22.24310067

**Authors:** Evan P. Troendle, Andrew J. Lee, Marina I. Reyne, Danielle M. Allen, Stephen J. Bridgett, Clara H. Radulescu, Michael Glenn, John-Paul Wilkins, Francesco Rubino, Behnam Firoozi Nejad, Cormac McSparron, Marc Niebel, Derek J. Fairley, Christopher J. Creevey, Jennifer M. McKinley, Timofey Skvortsov, Deirdre F. Gilpin, John W. McGrath, Connor G. G. Bamford, David A. Simpson

## Abstract

**Background:** Robust methods to track pathogens support public health surveillance. Both wastewater (WW) and individual whole genome sequencing (WGS) are used to assess viral variant diversity and spread. However, their relative performance and the information provided by each approach have not been sufficiently quantified. Therefore, we conducted a comparative evaluation using extensive individual and wastewater longitudinal SARS-CoV-2 WGS datasets in Northern Ireland (NI).

**Methods:** WGS of SARS-CoV-2 was performed on >4k WW samples and >23k individuals across NI from 14^th^ November 2021 to 11^th^ March 2023. SARS-CoV-2 RNA was amplified using the ARTIC nCov-2019 protocol and sequenced on an Illumina MiSeq. Wastewater data were analysed using Freyja to determine variant compositions, which were compared to individual data through time series and correlation analyses. Inter-programme agreements were evaluated by mean absolute error (MAE) calculations. WW treatment plant (WWTP) performances were ranked by mean MAE. Volatile periods were identified using numerical derivative analyses. Geospatial spreading patterns were determined by horizontal curve shifting.

**Findings:** Strong concordance was observed between wastewater and individual variant compositions and distributions, influenced by sequencing rate and variant diversity. Overall variant compositions derived from individual sequences and each WWTP were regionally clustered rather than dominated by local population size. Both individual and WW sequencing detected common nucleotide substitutions across many variants and complementary additional substitutions. Conserved spreading patterns were identified using both approaches.

**Interpretation:** Both individual and wastewater WGS effectively monitor SARS-CoV-2 variant dynamics. Combining these approaches enhances confidence in predicting the composition and spread of major variants, particularly with higher sequencing rates. Each method detects unique mutations, and their integration improves overall genome surveillance.

**Funding:** Individual sequencing was funded via the Belfast Health and Social Care Trust (Department of Health for Northern Ireland) and the COVID-19 Genomics UK (COG-UK) consortium, which was supported by the Medical Research Council (MRC), UK Research and Innovation (UKRI), the National Institute for Health Research (NIHR), the Department of Health and Social Care (DHSC), and the Wellcome Sanger Institute. The NI Wastewater Surveillance Programme was funded by the Department of Health for Northern Ireland. EPT was supported through the COG-UK Early Career Funding Scheme.

## INTRODUCTION

The COVID-19 pandemic presented unprecedented challenges to global societies, economies, and healthcare systems. Since its initial identification in late 2019^1^, the SARS-CoV-2 virus has continually mutated, generating new variants that spread globally, causing millions of infections and deaths^2^. This inherent capacity for rapid mutation and high transmissibility compelled the development of innovative strategies for surveillance, containment, and mitigation.

As SARS-CoV-2 mutations accumulate, genetic relationships between variants can be represented with a phylogenetic tree. Genomic surveillance samples sequences from the viral population and places them on the tree. This enables the classification of conserved mutation patterns, known as lineages. The Pango lineage classification system^3^ categorises global SARS-CoV-2 diversity. Assigning unique lineage labels enables researchers to identify transmission patterns^4^, monitor new variants, and assess control measures.

This classification system supports the variant of concern (VOC) framework for identifying and monitoring virus variants that pose significant public health risks. VOCs show evidence of increased transmissibility, severe disease, reduced treatment or vaccine effectiveness, or diagnostic failures. Identified variants undergo rigorous analysis to assess their public health impact. Genomic surveillance complements traditional epidemiology, providing critical evidence for informed decision-making by public health authorities to mitigate virus spread and protect public health.

Effective genomic surveillance depends on accurate variant identification, linked closely to sequencing effort, quality, and intensity^5^. During the pandemic, researchers and public health authorities used reverse transcription quantitative real-time polymerase chain reaction (RT-qPCR) and whole genome sequencing (WGS) to monitor SARS-CoV-2^6^. Global use of viral sequences tracked new variant emergence^7^. However, individual sequencing is resource-intensive and limited by potential uneven sampling and the typically low percentage of sequenced positive cases.

Wastewater (WW) screening for pathogens like poliovirus is long established in global surveillance^8,9^. Wastewater-based epidemiology (WBE) is based upon detection of viral RNA fragments shed in bodily secretions and excretions^10–12^ into municipal systems. Initially this was achieved using RT-qPCR, but the feasibility of wastewater-based sequencing (WBS) is now established^13–16^. Wastewater analysis anonymously surveys whole communities^17^, capturing symptomatic, asymptomatic, and pre-symptomatic infections^18,19^. This enhances understanding of community transmission and could serve as a source of early warning for outbreaks^20^.

WW contains a mixture of sequences, requiring computational deconvolution to determine the variants present. The widely used tool Freyja^21^ distinguishes and identifies variants in this complex mix by referencing genetic barcodes to perform depth-weighted demixing. Recent studies show alignment between Freyja and individual sequencing in tracking SARS-CoV-2 variants^18,22,23^.

The aims of this investigation are: 1) to quantify concordance between these approaches, examining the effects of sampling rate and variant diversity; 2) to evaluate how individual WW treatment plants (WWTPs) reflect the national picture or local diversity; 3) to determine the overlap in mutations detected by each method; and 4) to monitor intra-national transmission trends by detecting collective spreading patterns. To achieve these aims, we conducted a comparative evaluation of individual and wastewater-based WGS datasets in Northern Ireland (NI). By integrating geospatiotemporal metadata from both sequencing efforts and performing comparative analyses, we elucidate the synergy between these approaches and suggest how to combine them to improve surveillance.

## METHODOLOGY

For detailed methodologies, see the Supporting Information.

### WW sequencing programme

#### Wastewater treatment plant (WWTP) selection

31 WWTPs across NI were chosen based on population density, geographical distribution, and representation of urban and rural areas. This selection covers approximately 1,220,989 individuals, representing ~65% of NI’s population. See Table S1 for more information.

#### Sample collection

Composite WW samples were collected over 24-hour periods (Nov 14, 2021 - Mar 11, 2023) using an Isco Glacier autosampler.

#### Sample processing

Samples were clarified by centrifugation and concentrated using a CP-Select Concentrating Pipette™. Nucleic acids were extracted using the Roche MagNA Pure 96 Instrument.

#### SARS-CoV-2 quantitative reverse transcription polymerase chain reaction (RT-qPCR)

Extracted RNA was screened using AgPath-ID™ One-Step RT-qPCR Reagents and SARS-CoV-2 N1 + N2 Assay Kits on a LightCycler 480 II System.

#### Whole genome sequencing (WGS)

SARS-CoV-2 positive samples were sequenced using the Mini-XT SARS-CoV-2 protocol^24,25^ and ARTIC Network primers^26,27^. Libraries were prepared with Nextera XT and sequenced on an Illumina MiSeq. Sequencing FASTQs were generated onboard.

#### Quality control (QC)

Samples were excluded if they did not achieve ≥50% aligned whole-genome coverage with ≥10 bases per position. A timeline heatmap of the WW sample collection with QC inclusion/exclusion is shown in the SI as Figure S1.

### Individual sequencing programme

#### Individual Testing

Nasopharyngeal swabs underwent RT-qPCR testing, with positives sent for WGS. Pillar 1 testing targeted patients and frontline workers, while Pillar 2 involved the whole population until June 2022. See the Supporting Information, including Table S2 and Figure S2 for more information about the healthcare structure of NI.

#### WGS

From Nov 14, 2021, to Mar 11, 2023, 22,924 SARS-CoV-2 genomes were sequenced with Illumina and 556 with Nanopore. Using the Mini-XT protocol^24,25^, cDNA was amplified by tiled PCR with ARTIC primers^26,27^, purified, quantified, and sequenced on Illumina MiSeq or Oxford Nanopore platforms. Sequencing FASTQs were generated onboard.

#### Quality control (QC)

Samples were excluded if they lacked sufficient collection time and location metadata, or if they failed to achieve either ≥50% whole-genome coverage (≥10 reads per base for Illumina or ≥20 for Nanopore) or a ≥10,000 base continuous sequence without Ns (i.e., ambiguous or missing nucleotide bases).

### Geographic Information Systems (GIS)

#### Geospatial Mapping

Geopandas v0.14.3 was used for geographic visualisations and analyses.

#### Local Government Districts (LGDs) in NI

NI comprises 11 LGDs, responsible for local government. A shapefile from OpenDataNI was used for LGD-level geospatial analysis. See the Supporting Information and Table S3 for additional information.

### Bioinformatics

#### Nextflow pipeline

Bioinformatics analyses were conducted using the Illumina Nextflow pipeline developed by the ARTIC network, tailored for SARS-CoV-2 data processing. The pipeline, which automates fieldbioinformatics tools, was utilised in our modified version from https://github.com/QUB-Simpson-lab/ncov2019-artic-nf. Software updates included SAMtools^28^ and BCFtools^29^ upgraded to v1.18, trim_galore (https://github.com/FelixKrueger/TrimGalore) to v0.6.10, and iVar^30^ to v1.4.2. Additionally, Freyja^21^ v1.4.9 for variant calling and Pangolin^31^ v4.3.1 for lineage calling were integrated.

The workflow involved several key steps: The SARS-CoV-2 reference genome^1^ (i.e., NC_045512.2, MN908947.3, or hCoV-19/Wuhan/WIV04/2019) and ARTIC primer schemes were downloaded and indexed using BWA^32^. Paired-end FASTQ files were pre-processed with trim_galore for consistent adapter and quality trimming. Reads were mapped to the reference genome using BWA mem, and primer sequences were removed with iVar trim. Consensus sequences were generated using iVar consensus with SAMtools mpileup output, and variants were called using iVar variants and Freyja variants. Lineage calling was performed using Pangolin, and Freyja demix was used for depth-weighted demixing with a depth cutoff of 10. A flowchart of the pipeline can be found in the Supporting Information as Figure S3.

#### Classification of SARS-CoV-2 variants into mutation constellations

We used regular expression patterns to parse and categorise Pango lineage names into mutation constellations, which are collections of functionally significant mutations arising independently in the virus’s genome. This method organised Pango lineages demixed by Freyja and identified by Pangolin (see Table S4). Our classifications have subtle variations from those curated by the Pango Network (https://cov-lineages.org/constellations.html), aligning more closely with the variant mix detected in NI.

#### Geospatial abundance time series calculations of SARS-CoV-2 constellations

Using pandas v2.2.2, we estimated SARS-CoV-2 constellation abundance across Northern Ireland, its LGDs, and the WWTP catchment areas. Anonymised sample time of collection and LGD metadata were obtained from CLIMB (COG-UK) (and is also available on GISAID^7^). Each sample was mapped to a constellation (individuals) or constellations (WW), and daily counts (individuals) or means (WW) were computed, ensuring continuous time series with interpolation. A 15-day centred rolling window smoothed fluctuations, and percentages normalised data for comparisons.

#### Calculation of mean absolute errors (MAEs)

Circulating mutation constellations were defined as those with an abundance of ≥ 10%. Daily mean absolute error (MAE) for each of these constellations was computed by summing the absolute differences between corresponding constellation abundances in their time series and dividing by the number of circulating variants.

#### Quantitative determination of volatile periods

We used numerical derivative analysis to identify periods of abundance fluctuations within each programme. Smoothed (7-day centred rolling window average) daily slopes of the abundance time series were calculated, and net changes in constellation abundances were determined by summing the absolute values of these derivatives. Volatile periods were identified using a net 2%/day threshold, marking those exceeding it as volatile.

#### Correlations

Pearson correlation coefficients and associated p-values were calculated using SciPy^33^ 1.13.0 (*stats.pearsonr*).

#### WWTP quantitative rankings

To evaluate the reflectivity of wastewater treatment plants (WWTPs) in detecting SARS-CoV-2 variants, we compared variant abundances from individual population sequencing (Pillar 2) with those from each WWTP. Mean absolute error (MAE) time series per WWTP versus Pillar 2 samples in NI were used to quantify mean discrepancies between the surveillance methods. Ranking based on mean MAE assesses agreement over time, identifying the most and least reflective WWTP sites for SARS-CoV-2 variant detection.

#### Assessing complementarity between programmes using the Freyja UShER substitution barcode database

We used the Freyja UShER substitution barcode database to categorise nucleotide substitutions in individual and wastewater samples. This matrix-format database assigns columns to genomic substitutions (e.g., G210T) aligned to the reference genome and rows to unique SARS-CoV-2 Pango lineages, denoted by binary values indicating substitution presence. Each lineage’s ‘barcode’ serves as a genetic fingerprint, providing the set of expected present substitutions.

For each constellation, substitutions are categorised into three groups: ‘core’ substitutions found in all lineage barcodes, ‘accessory’ substitutions present in some but not all barcodes within the constellation, and ‘other’ observed substitutions not catalogued by Freyja.

We compared the number and identity of substitutions to assess how well the two sequencing programmes complement each other across various geospatial scales: NI-wide, by LGD, and by WWTP. Substitutions were detected in individual and WW samples using minimum allele frequencies of 0.75 and 0.25, respectively, with a minimum read depth of 10 from Freyja/iVar outputs.

#### Customisation of Freyja UShER barcodes

During transitions between mutation constellations (e.g., Delta-like to Omicron BA.1-like), inaccuracies in reported abundances occurred, likely due to Freyja encountering challenges in distinguishing between mixed samples of parent lineages and their recombinants. To resolve this, we customised the Freyja UShER substitution barcodes database to exclude recombinant Pango lineages (X*) while retaining XBB* lineages, recovering accurate abundance estimation within relevant mutation constellations. Further details are available in the Supporting Information (see Figure S4).

## RESULTS

### Variant abundances derived from WW match individual sequencing in Northern Ireland

Samples were collected from individuals present or living within Northern Ireland (NI) as well as from wastewater treatment plants (WWTPs) (Figure 1, left). Abundance time series were calculated and used to visualise the prevalence and distribution of each SARS-CoV-2 mutation constellation across NI (see *Methodology*). Comparison of abundance time series from WW surveillance with those from individual sequencing programmes (Figure 1, right) demonstrates excellent agreement, underscoring the efficacy of the recently introduced wastewater-based sequencing (WBS) programme in NI.

**Figure 1.**
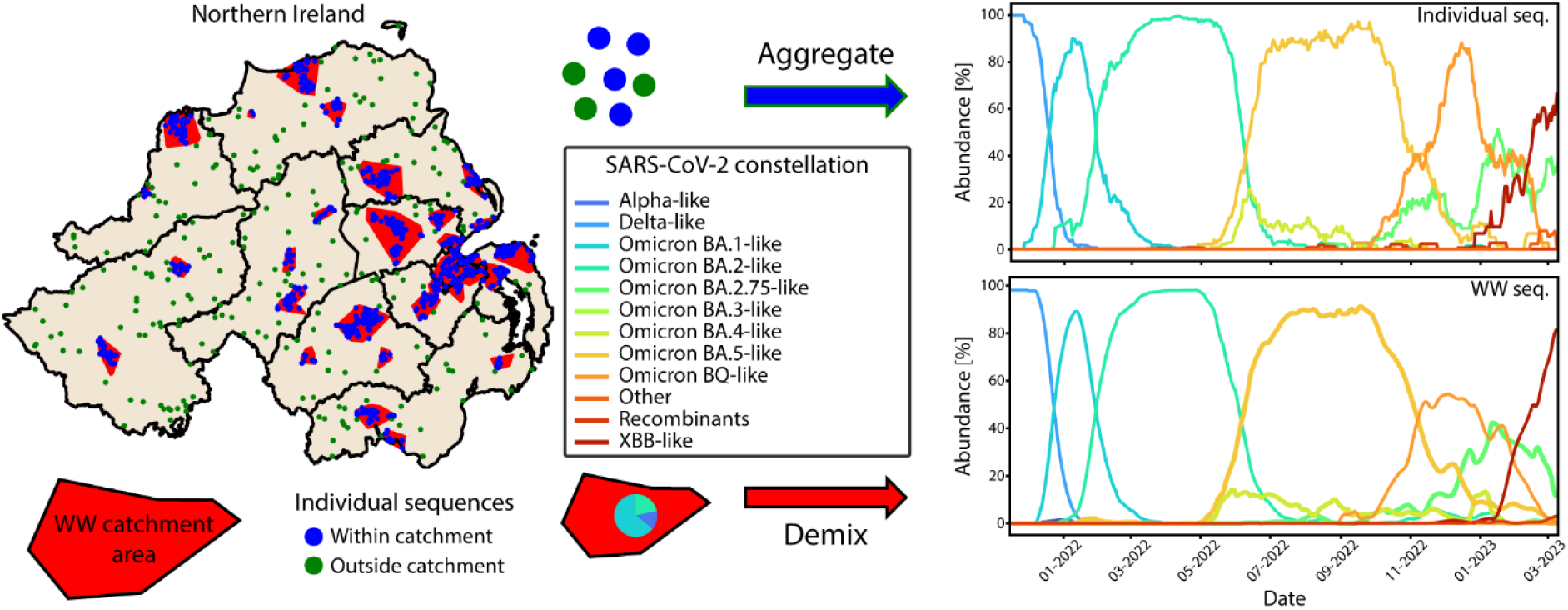
Surveillance of SARS-CoV-2 in Northern Ireland: Sampling strategy and temporal dynamics. Left: *Spatial Distribution of Sequences and Wastewater Treatment Plant Catchment Areas* – A map of Northern Ireland (NI) displays the boundaries of the 11 local government districts with wastewater treatment plant catchment areas highlighted in red and illustrative individual samples either within (blue dots) or outside them (green dots). Right: *Ensemble Average SARS-CoV-2 Constellation Abundances* – The ensemble average SARS-CoV-2 constellations calculated across the entirety of NI as determined by aggregating individual sequences and demixing wastewater sequences, respectively.

### Sources of variation between wastewater and individual sequencing

The SARS-CoV-2 constellation abundances defined from each sequencing programme did exhibit some variability (Figure 2A); visual inspection reveals periods of both stronger and weaker agreements between individual and WW sequencing. Mean absolute error (MAE) time series (Figure 2B) clearly indicated pronounced disagreements during transitions between dominant constellations and periods with multiple circulating constellations. To quantify the volatility in abundances, we used numerical derivative analysis to determine daily curve slopes (Figure 2C) and calculated net changes in constellation abundances (Figure 2D). Volatile periods were defined as those exceeding a net 2%/day threshold.

**Figure 2.**
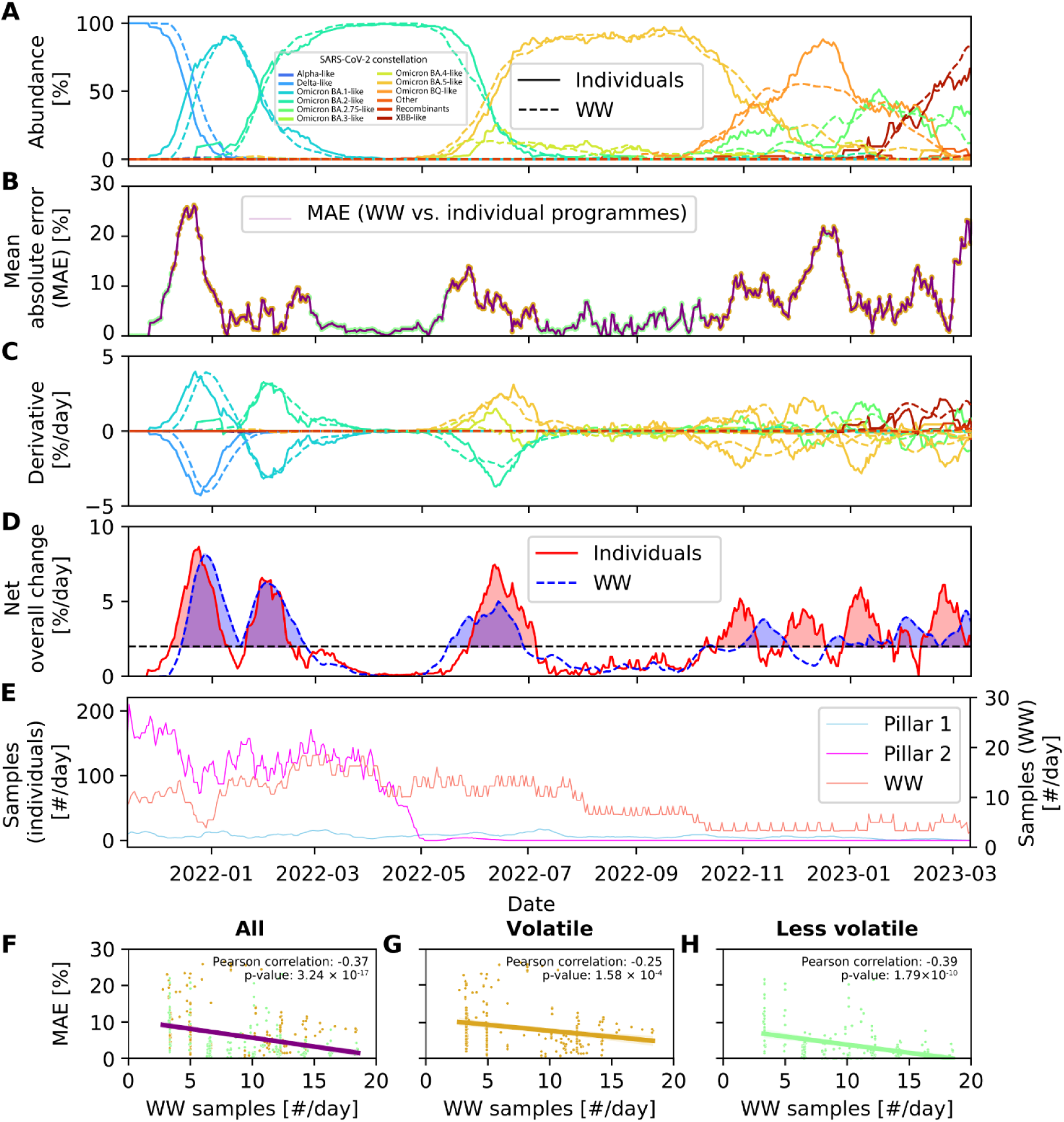
Comparative Analysis of Overall SARS-CoV-2 Abundance and Sampling Rates in Northern Ireland. A: *Ensemble SARS-CoV-2 constellation abundances comparison* – This panel presents the SARS-CoV-2 constellation time series from Figure 1, facilitating visual comparison. Solid lines represent abundances from individual sequencing programmes, while dashed lines depict those from the wastewater (WW) sequencing programme. B: *Quantification of temporal deviation between sequencing programmes* – The purple line illustrates the daily mean absolute errors (MAEs) between all constellations observed in WW-based sequencing and those in individual-based sequencing programmes. Points labelled in goldenrod align with transitional periods detected from the wastewater programme, while points labelled in pale green indicate more stable periods. C: *Rates of change in ensemble SARS-CoV-2 constellation abundances* – The numerical derivative was calculated for each time series in panel A, yielding daily curve slopes. D: *Net overall changes in constellation abundances to determine transition periods* – The solid red line represents the daily sums of absolute changes for individual sequencing programmes, and the dotted blue line represents the wastewater sequencing programme from C. A horizontal dotted black line indicates a 2%/day threshold, with periods exceeding this threshold shaded in red and blue. E: *Sampling rates of Pillar 1, Pillar 2, and WW sequencing programmes* – Pillar 1 sampling rates (#/day) are shown in cyan, Pillar 2 in magenta, and WW samples on the secondary y-axis in salmon. F-H: *Correlations between WW sequencing rates and inter-programme agreement* – These scatter plots depict correlations between all WW sequencing rates (B) and mean absolute errors (E), representing inter-programme agreement. Points within transition periods are labelled goldenrod (G), while those outside of transition periods are labelled pale green (H).

Comparing MAEs between programmes (Figure 2B) with sequencing rates (Figure 2E) revealed decreased agreement as sequencing rates declined, especially during lineage transitions and heightened SARS-CoV-2 variant co-circulation. Correlation analyses confirmed an inverse relationship between wastewater sequencing rates and MAEs, with a negative correlation (Pearson r = −0.37, p-value = 3.24 × 10^−17^) (Figure 2F). The correlation remained (Pearson r = −0.25, p-value = 1.58 × 10^−4^) during volatile periods (Figure 2G), but strengthened (Pearson r = −0.39, p-value = 1.79 × 10^−10^) during less volatile periods (Figure 2H). During these volatile periods, the regression line (Figure 2G) shows a higher y-intercept and a flatter slope, implying that reducing Mean Absolute Error (MAE) requires more sequencing compared to less volatile periods at similar levels. Further analysis of MAE distribution characteristics reveals variability ranging from 0.03% to 26.15%, with a mean MAE of 6.19% and a standard deviation of 5.69%.

### Assessing WWTP performance for matching individual SARS-CoV-2 surveillance

Comparison of variant abundances detected through public sequencing (Pillar 2) and by each WWTP revealed varied agreement across NI sampling sites (Figure 3A). Per-WWTP mean absolute error (MAE) time series were computed to assess the level of disagreement between the two methods, as shown by Newtownbreda (NTB), Belfast (BEL), and Enniskillen (ENN) in Figure 3B. Lower mean MAE values indicated higher agreement, while higher values indicated greater disparities. Sorting these mean MAE values in ascending order (Figure 3C) provided a reliability metric to rank the performance of the sites in reflecting the overall NI SARS-CoV-2 variant detection.

**Figure 3.**
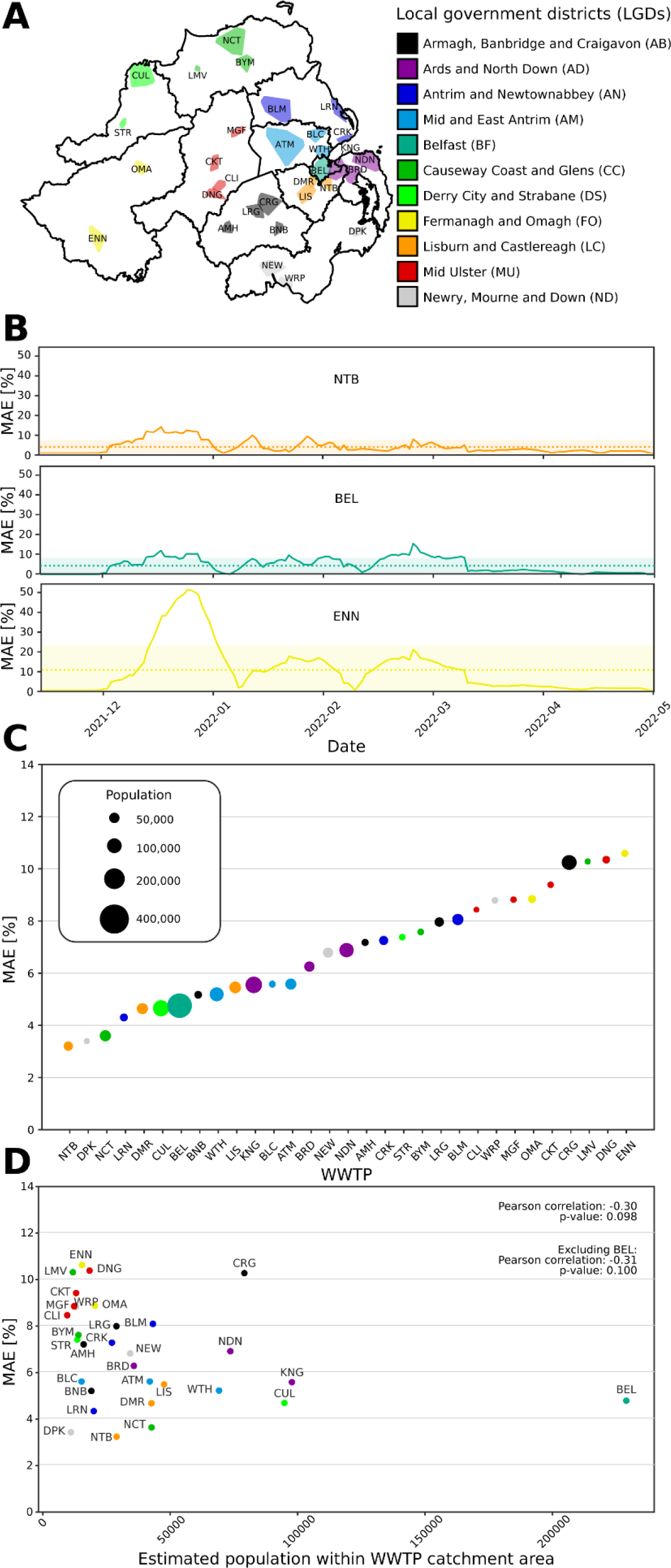
Performance evaluation of the WWTPs in comparison to Pillar 2 sequences. A: Annotated map of WWTP catchment areas in NI, coloured by local government district (LGD) and labelled with abbreviations: ATM: Antrim, AMH: Armagh, BLC: Ballyclare, BLM: Ballymena, BYM: Ballymoney, BRD: Ballyrickard, BNB: Banbridge, BEL: Belfast, CRK: Carrickfergus, CLI: Coalisland, CKT: Cookstown, CRG: Craigavon, CUL: Culmore, DPK: Downpatrick, DNG: Dungannon, DMR: Dunmurry, ENN: Enniskillen, KNG: Kinnegar, LRN: Larne, LMV: Limavady, LIS: Lisburn, LRG: Lurgan, MGF: Magherafelt, NEW: Newry, NTB: Newtownbreda, NCT: North Coast, NDN: North Down, OMA: Omagh, STR: Strabane, WRP: Warrenpoint, and WTH: Whitehouse. B: Per-WWTP mean absolute error (MAE) time series, with NTB, BEL, and ENN shown as examples. The mean value is dotted, and the standard deviation is shaded with semi-opacity. C: Relative ranking of WWTPs by mean MAE, presented in a scatter plot with point size indicating estimated population within each WWTP catchment area. D: Correlation between mean MAE and estimated population within the catchment, displayed as a scatter plot with correlation results.

Newtownbreda (NTB), Downpatrick (DPK), North Coast (NCT), Larne (LRN), Dunmurry (DMR), Culmore (CUL), Belfast (BEL), Banbridge (BNB), Whitehouse (WTH), Lisburn (LIS), and Kinnegar (KNG) emerged as the 10 sites with the lowest mean MAEs. While these sites tracked the sampled NI population well, lower-ranking sites (e.g., Enniskillen (ENN), Dungannon (DNG), Limavady (LMV), Craigavon (CRG), Cookstown (CKT), Omagh (OMA), Magherafelt (MGF), Warrenpoint (WRP), Coalisland (CLI), and Ballymena (BLM)) likely reflect distinct local viral dynamics that were not captured by Pillar 2 data. This underscores the additional insights these sites can provide, enhancing our understanding of viral dynamics in NI.

Additionally, the Pearson coefficient of −0.30 (p-value = 0.098) for the correlation between estimated population coverage and mean MAE (Figure 3D) suggests that the largest population centres should not necessarily be prioritised for WWTP sentinel sites and invites further consideration of the role of smaller sites.

### Complementarity between wastewater and individual WGS for SARS-CoV-2 mutation detection

Distinct patterns of nucleotide substitution are detected by wastewater and individual sequencing programmes, both at the national and local government district level. We categorised substitutions into three groups: ‘core’ substitutions, which are highly conserved and define mutation constellations (e.g., common substitutions in Delta-like, BA.1-like, and XBB-like lineages); ‘accessory’ substitutions, which are lineage-specific but not universally conserved within mutation constellations; and ‘other’ substitutions, which are not captured in the Freyja UShER substitution barcode database and may represent individual-specific mutations or local infection clusters (see Figure 4A).

**Figure 4.**
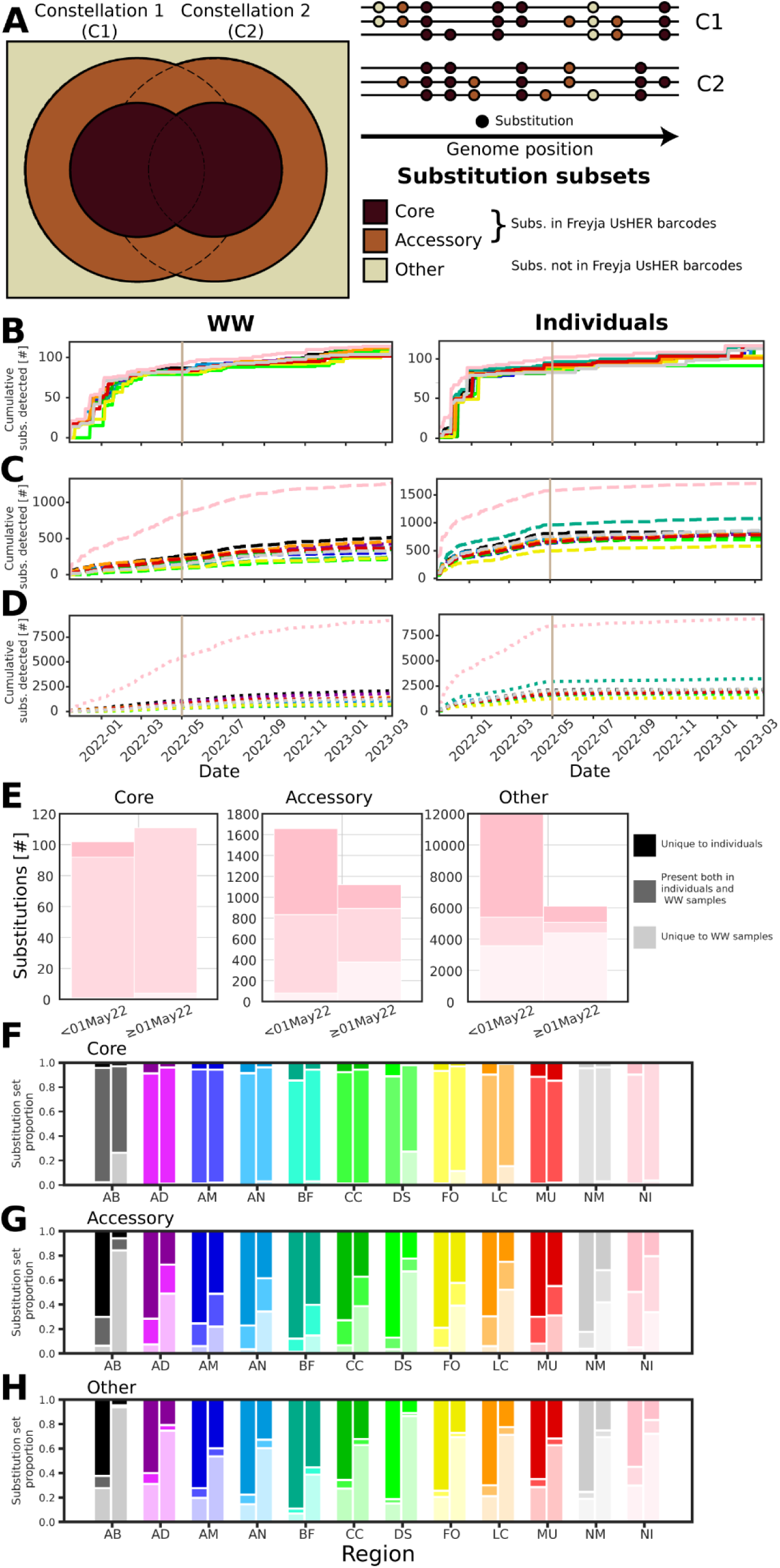
Complementarity of individual and WW WGS. A. *Schematics of the substitution classification system.* Left: concentric Venn diagram showing intersections of substitution sets. The inner Venn diagram (chocolate) represents ‘core’ substitutions found in every lineage within a mutation constellation. The outer Venn diagram (rust brown) shows ‘accessory’ substitutions present in some but not all lineages. The area outside the Venn diagrams (vanilla) represents ‘other’ substitutions not included in the Freyja UShER barcodes. Upper right: diagram of multiple lineage genomes grouped into constellations with substitutions coloured according to their classification in the Venn diagram. **B-D**. Newly detected core, accessory, and other substitutions over time in wastewater (WW) on the left and individuals on the right, shown per Local Government District (LGD) and for all NI samples. Vertical tan lines mark May 1, 2022, when Pillar 2 sampling decreased sharply towards zero. **E**. Set sizes of detected core, accessory, and other substitutions, showing unique detections and overlaps before and after the decline in Pillar 2 sampling. **F-H**. Proportional overlaps and unique detections of core, accessory, and other substitutions per LGD and for all NI samples.

We tracked the emergence of substitutions over time in wastewater samples and within individual sequencing data (see *Methodology*). Core substitutions were consistently captured by both sequencing approaches (see Figure 4B&F), demonstrating their reliability in identifying mutation constellation-defining mutations. However, there was more variability in the accessory and other substitutions detected, even when using more stringent detection thresholds. The numbers detected by only one approach increased from very few ‘core,’ to more ‘accessory’ to a majority of ‘other’ substitutions. The set sizes and proportions detected in each case are presented in Table S5.

### Temporal shifting reveals conserved geospatial patterns of SARS-CoV-2 spread in Northern Ireland

Focusing on the BA.1-like→BA.2-like (excluding BA.2.75-like) transition, a consistent pattern in the relative arrival times of BA.2-like variants emerged. Newry, Mourne and Down (ND), Armagh, Banbridge and Craigavon (AB), and Lisburn and Castlereagh (LC) led in early arrivals, while Causeway Coast and Glens (CC), Derry City and Strabane (DS), and Fermanagh and Omagh (FO) showed later arrivals (Figure 5A).

**Figure 5.**
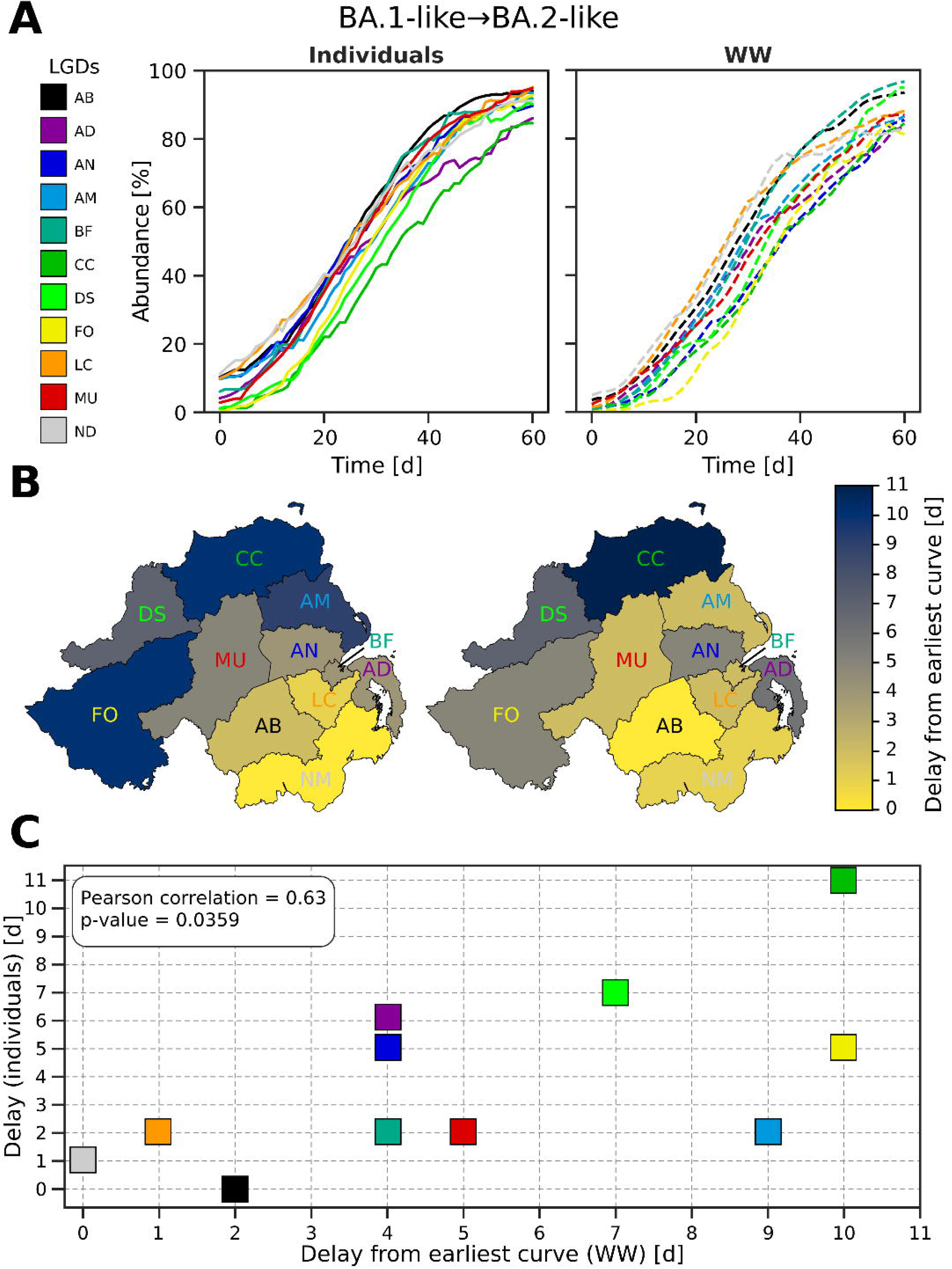
Detecting apparent geospatial spreading patterns through temporal curve shifting. A. The rise of BA.2-like variants over 60 days during the BA.1-like to BA.2-like (excluding BA.2.75-like) transition (Jan 3, 2022 – Mar 4, 2022) for individuals (solid lines, left) and wastewater (dotted lines, right). Local Government Districts (LGDs) are colour-coded and abbreviated per the Troendle-Rice-Simpson-Skvortsov scheme (see Figure 3A and Table S3). B. Choropleth maps showing delay times derived from A, with bright yellow for the earliest and dark navy for the latest observances. C. Correlation scatter plot of geospatial spreading delays, comparing individuals (y-axis) and wastewater (x-axis), with Pearson correlation coefficient and p-value annotated.

Visualisations suggest that the Belfast area and southern regions (e.g., LC, AB, ND) detected new variants earlier, whereas northern and western regions (e.g., CC, FO, DS) lagged (Figure 5B). Mid and East Antrim (AM), Mid Ulster (MU), and FO exhibited discrepancies, with individual sequencing detecting variants earlier than wastewater surveillance in these regions.

The correlation between detection dates (Pearson r = 0.63, p-value = 0.0359) indicates decent agreement between individual and WW data (Figure 5C). To see additional animations depicting spreading and detection patterns observed from WW and in individuals, please refer to the Supporting Information.

## DISCUSSION

The closely matching abundances of SARS-CoV-2 mutation constellations detected from WW and individual sequencing in NI demonstrate the efficacy of both programmes. Both approaches also showed consistent variant geospatial spreading patterns.

Despite these independent sequencing approaches each having inherent biases, as sequencing levels increase their findings converge towards what is likely to be the true picture of pathogen dynamics. Comparison of their similarity therefore provides an objective assessment of performance. This assessment should be based upon volatile periods with rapid changes in variant abundances because less sequencing is required to achieve concordance during periods of low co-circulation, when a predominant variant is present (see Figure 2 (cf. F-H)). Our analysis enables unbiased detection of these volatile periods.

Any genome surveillance programme must consider the resources required to collect, process and sequence samples. Our findings contribute to this decision-making process by determining the minimum necessary level to achieve target mean absolute errors (MAEs) between individual samples and wastewater results. This analysis strategy can be applied globally to provide evidence guiding the refinement of other programmes and assess effective sequencing strategies. Aiming to keep a MAE under 10% provides confidence that the strategy employed is accurately capturing pathogen dynamics. For the NI WBS programme, achieving a 10% MAE required approximately 15 WWTPs to be sampled on average every day during volatile periods, while reducing the MAE to 5% might necessitate approximately 20.

Individual- and wastewater-based sequencing methods present distinct advantages and limitations for pathogen surveillance. WBS cannot capture samples from populations residing in rural areas outside municipal water systems, potentially underrepresenting certain demographics (See Figure 1). Nonetheless, the broad community-level trends captured by WBS may not be as accurately measured by individual sequencing due to sampling limitations, such as over-representation of healthcare-related cases. Individual sequencing generates largely complete viral genomes, whereas WBS sequences are fragmented and genomes are not directly attributable to individuals. Wastewater surveillance detects viral shedding from asymptomatic and presymptomatic individuals unlikely to be individually sampled. While the information contributed by each programme correlates with its sequencing rate (see Figure 4), WBS provided substantial additional insights even during periods of high individual sequencing rates. The differing strengths and weaknesses underscore the importance of integrating both approaches to achieve a more comprehensive understanding of viral diversity and transmission dynamics. For the cost of collecting and processing a relatively small number of WW samples, a dedicated WBS surveillance programme could provide systematic population-level monitoring. Nonetheless, individual sequencing may capitalise on existing healthcare and research infrastructures, provides additional detailed information, and is essential for elucidation of outbreak transmission trees.

The weaker than expected correlation between population size and the national SARS-CoV-2 variant landscape challenges the assumption that larger population WWTPs should always be prioritised for surveillance. Strategic selection of sentinel sites for WBS is required to reflect both global and local patterns. The extent to which each WWTP site matched pathogen mutations detected in individuals varied over time (Figure 3), influenced by the viral dynamics within the WWTP catchment area. Our findings demonstrate that our WBS approach was sufficiently balanced spatially and temporally to address the study objectives.

Figure 5 illustrates the coinciding geospatiotemporal patterns of pathogen spread observed through both WBS and individual WGS programmes. Although the patterns were less similar in other waves with lower sequencing rates, these findings underscore the value of integrating both datasets to enhance the granularity and accuracy of epidemiological monitoring. By capturing regional variations in pathogen dynamics, localised surveillance strategies can be more effectively tailored to address specific areas of concern.

Given the ability of WBS to add information, integration of other environmental sampling approaches such as air, surface, water, wildlife, and food/agriculture sampling could further enhance surveillance. Air sampling captures viral particles in aerosols, while surfaces in public spaces and healthcare settings serve as reservoirs for viral RNA. Water sampling from natural bodies could reveal environmental transmission routes, while monitoring wildlife and food/agriculture environments could identify potential spillover events and routes of viral contamination. These methods collectively provide insights into community-level transmission dynamics, identify virus hotspots, and trace transmission chains in healthcare settings, thereby enhancing our understanding of viral circulation and transmission pathways alongside current genomic surveillance efforts based on wastewater and individual sampling.

WW genome surveillance faces significant analytical challenges, primarily related to the complex task of demixing multiple co-circulating viral lineages. This process in Freyja entails deciphering signals influenced by sequencing depth and associated mutational frequencies relative to curated barcodes. The inherent combinatorial complexity of genomic mutations and the diversity observed in viral genomes further complicate efficient data analysis. Differentiating between similar barcodes is particularly challenging, especially when specific mutations present or absent in individual samples can confound demixing efforts. Distinguishing between a mixture of parent lineages and recombinant strains also adds complexity.

Mutation detection thresholds from WBS influence the overlap of mutation profiles observed between methods. Setting appropriate detection thresholds is crucial to extract meaningful information from mixed wastewater samples, considering factors such as sequencing depth, sequencer quality scores, and environmental conditions. Our study demonstrates that our selected thresholds (allele frequency ≥ 0.25, depth ≥ 10) effectively capture core substitutions, enhancing confidence in the veracity of detected accessory and other substitution sets, supporting our approach despite the absence of a definitive ground truth. Additionally, wastewater-based surveillance effectiveness varies by location due to environmental factors like chemical composition, local biota, and climate conditions^34^. highlighting the complexity in accurately tracing respiratory virus spread and evolution through wastewater surveillance. Continued advancements in analytical technologies and techniques will help to address these complexities.

Moving forward, reconstructing genomes by inferring and stitching together individual reads based on their sets of mutations offers a promising approach to overcome some of the challenges associated with current demixing algorithms. Rather than relying on *a posteriori* mutation barcodes for demixing, *de novo* genome assembly reconstructs viral genomes directly from fragmented reads. By analysing these assembled sequences, researchers could gain more accurate insights into the genetic diversity and evolutionary trajectories of metagenomic populations present in their wastewater samples. This approach would shift the focus from simply detecting the presence of individual mutations of a target genome in the mix to understanding the genomic context in which the mutations occur, potentially enhancing the breadth, accuracy, and reliability of wastewater-based epidemiological studies.

## CONCLUSIONS

Our study has rigorously quantified the concordance between various sequencing approaches across multiple spatial and temporal scales. Evaluation of the alignment of individual wastewater treatment plants (WWTPs) with national trends revealed nuanced insights into the contribution of each site to the larger surveillance network. Larger sites do not always provide the most representative data, highlighting the limitation of relying on a single site to adequately capture regional diversity. Analysis of mutation overlap across methods enabled establishment of effective detection parameters and strategies, crucial for enhancing the accuracy of variant identification.

A key finding is that the extensive spatial coverage and the low cost per individual represented by wastewater-based sequencing (WBS) complement the capabilities of individual sequencing efforts. Both approaches contribute to elucidation of intra-national transmission trends and collective spreading patterns. We therefore recommend the integration of WBS within existing genomic surveillance frameworks to improve pathogen monitoring by: 1) expanding the use of WBS to localities currently lacking comprehensive genomic surveillance; 2) ensuring a balanced approach to geographic and temporal resolution in WBS implementation; and 3) adopting a dynamic resource allocation strategy to respond swiftly to emerging threats, such as increasing sample collections in nearby WWTPs upon detection of a new variant. These measures will collectively enhance the capacity to detect, monitor, and respond to infectious disease threats at local and national levels, supporting more effective public health interventions.

In summary, our comparative and quantitative evaluation demonstrates the complementary roles of WBS and individual WGS in enhancing our understanding of SARS-CoV-2 viral diversity and dynamics.

## Data Availability

All data produced in the present study are available upon reasonable request to the authors.

https://github.com/QUB-Simpson-lab/ncov2019-artic-nf

## ACKNOWLEDGMENTS

We would like to thank: Jonathon D. Coey, Pearce Allingham, Ashley Levickas, Stephen H. Bell, Jonathan Lock, and Aaron G. Canton Bastarrachea, for support within the Queen’s University Belfast (QUB) School of Biological Sciences (SBS) and Wastewater Surveillance Laboratory; members of the QUB COVID-19 Genomics (QUB-COG) and QUB Genomics Core Technology Unit (QUB-GCTU) teams, including Marc-Aurel Fuchs, Miao Tang, Arun Mahesh, Deborah Lavin, Syed Umbreen, Alan M. Rice, Arthur Fitzgerald, Mark Smyth, Eilís McColgan, Zoltan Molnar, Chris Baxter, Aditi Singh, Fiona Rogan, Julia Miskelly, Sarah Sonner, and Courtney Ward; members of the Regional Virus Laboratory (RVL) Belfast Health and Social Care Trust (BHSCT) team, including James P. McKenna, Conall McCaughey, Tanya Curran, Susan Feeney, Alison Watt, Ciara Cox, and Mairead Connor. All named individuals above helped with various aspects of our SARS-CoV-2 individual and wastewater genomics sample collection, processing, sequencing, metadata curation, and bioinformatic analyses. We would also like to thank Declan T. Bradley of the NI Public Health Agency (PHA) and the QUB Centre for Public Health (CPH) for several helpful discussions. Lastly, we would like to thank the now-dissolved COG-UK consortium for their lasting contributions to supporting global genomic surveillance efforts.

## AUTHOR CONTRIBUTIONS

EPT, JWM, CGGB, and DAS designed the study. EPT and DAS drafted the manuscript. EPT performed depth-weighted demixing of Northern Ireland (NI) wastewater (WW) samples with Freyja. EPT also developed downstream analysis algorithms, including managing and extending the fork of the ncov2019-artic-nf pipeline, performing quality control for sample exclusion, classifying SARS-CoV-2 mutation constellations, calculating geospatiotemporal constellation abundance, sequencing rate, and mean absolute error (MAE) time series, assessing WWTP site performance and ranking, evaluating complementarity by substitution categorisation and accumulation, identifying statistical correlations between variables, and detecting geospatial spreading patterns by shifting time series. EPT also produced associated data and manuscript figures. AJL, MIR, and DMA conducted extensive WW sample collection, processing, and sequencing. SJB and EPT performed ncov2019-artic-nf bioinformatics processing of individual and WW samples. SJB directed individual sample uploads to CLIMB. CHR and MG conducted extensive individual and WW RT-qPCR, library preparation, and sequencing to support the project. EPT, JP-W, FR, and CJC contributed bioinformatic analyses of the NI WW data. EPT, BFN, CM, and JMM contributed to geospatial analysis of NI WW and individual sequencing data. MN and DJF contributed contextual analysis from a public health perspective. DAS, TS, DFG, CGGB, and JWM managed the integration of the individual and WW sequencing and genome surveillance programmes. All authors (EPT, AJL, MIR, DMA, SJB, CHR, MG, J-PW, FR, BFN, CM, MN, DJF, CJC, JMM, TS, DFG, JWM, CGGB, and DAS) contributed to editing the manuscript, performing literature review, and writing the introduction, methodology, results, discussion, and conclusions. EPT, JWM, and DAS secured funding supporting the research.

