## Supporting Information for "Combining individual and wastewater whole genome sequencing improves SARS-CoV-2 surveillance"

#### TITLE:

### Supporting Information for "Combining individual and wastewater whole genome sequencing improves SARS-CoV-2 surveillance"

#### AUTHOR LIST:

Evan P. Troendle[1,*], Andrew J. Lee[2], Marina I. Reyne[2], Danielle M. Allen[2], Stephen J. Bridgett[1], Clara H. Radulescu[1], Michael Glenn[1], John-Paul Wilkins[2], Francesco Rubino[2], Behnam Firoozi Nejad[3], Cormac McSparron[3], Marc Niebel[4], Derek J. Fairley[4], Christopher J. Creevey[2,5], Jennifer M. McKinley[3], Timofey Skvortsov[6], Deirdre F. Gilpin[6], John W. McGrath[2,5], Connor G. G. Bamford[2,5,*], and David A. Simpson[1,*]

#### AFFILIATIONS:

[1]. Wellcome-Wolfson Institute for Experimental Medicine, School of Medicine, Dentistry and Biomedical Sciences, Queen's University Belfast, Belfast, Northern Ireland, United Kingdom, BT9 7BL

[2]. School of Biological Sciences, Queen's University Belfast, Belfast, Northern Ireland, United Kingdom, BT9 5DL

[3]. Geography, School of Natural and Built Environment, Queen's University Belfast, Belfast, Northern Ireland, United Kingdom, BT9 6AZ

[4]. Regional Virology Laboratory, Belfast Health and Social Care Trust, Royal Victoria Hospital, Belfast, Northern Ireland, United Kingdom, BT12 6BA

[5]. Institute for Global Food Security, Queen's University Belfast, Belfast, Northern Ireland, United Kingdom, BT9 5DL

[6]. School of Pharmacy, Queen's University Belfast, Belfast, Northern Ireland, United Kingdom, BT9 7BL

Wastewater surveillance shows both early detection of substitutions associated with emerging constellations and prolonged presence of substitutions from fading constellations 15

Figure S5. Set-based substitution tracking of SARS-CoV-2 constellation transitions 17

Quantitative assessment of temporal shifts between WW and Pillar 2 sequencing in capturing the rise of SARS-CoV-2 constellations 18

Figure S6. Temporal deviations and optimal alignment analysis between WW and Pillar 2 (individual) sequencing 19

Substitution sets tracked in Figure S5 20

Table S6. Substitution sets tracked in transitions (see Figure S5). 20

Figure S7. Coverage of ARTIC V4 amplicon primers for exemplary WW WGS. 21

Figure S8. Genome coverage for an exemplary subset of the WW WGS. 21

References: 22

#### EXTENDED METHODOLOGY:

##### WW sequencing programme

Wastewater treatment plants (WWTPs)

**Table S1. Wastewater treatment plants (WWTPs)**

| WWTP Name | Abbreviation | Samples [#] | Population coverage | Catchment area [km$^2$] | Local Government District |
|---|---|---|---|---|---|
| **Antrim** | ATM | 147 | 41735 | 28.26 | Antrim and Newtonabbey |
| **Armagh** | AMH | 147 | 15749 | 10.11 | Armagh City, Banbridge and Craigavon |
| **Ballyclare** | BLC | 94 | 14933 | 7.97 | Antrim and Newtonabbey |
| **Ballymena** | BLM | 91 | 42969 | 27.00 | Mid and East Antrim |
| **Ballymoney** | BYM | 144 | 13721 | 8.39 | Causeway Coast and Glens |
| **Ballyrickard** | BRD | 148 | 35497 | 14.94 | Ards and North Down |
| **Banbridge** | BNB | 151 | 18803 | 8.32 | Armagh City, Banbridge and Craigavon |
| **Belfast** | BEL | 150 | 228939 | 65.00 | Belfast |
| **Carrickfergus** | CRK | 100 | 26860 | 11.68 | Mid and East Antrim |
| **Coalisland** | CLI | 102 | 9312 | 7.36 | Mid Ulster |
| **Cookstown** | CKT | 148 | 12797 | 8.70 | Mid Ulster |
| **Craigavon** | CRG | 148 | 78899 | 49.07 | Armagh City, Banbridge and Craigavon |
| **Culmore** | CUL | 136 | 94655 | 48.28 | Derry City and Strabane |
| **Downpatrick** | DPK | 118 | 10755 | 6.24 | Newry, Mourne and Down |
| **Dungannon** | DNG | 149 | 18079 | 9.78 | Mid Ulster |
| **Dunmurry** | DMR | 152 | 42397 | 14.38 | Lisburn and Castlereagh |
| **Enniskillen** | ENN | 138 | 15115 | 23.34 | Fermanagh and Omagh |
| **Kinnegar** | KNG | 149 | 97582 | 35.64 | Ards and North Down |
| **Larne** | LRN | 126 | 19724 | 11.10 | Mid and East Antrim |
| **Limavady** | LMV | 136 | 11538 | 4.81 | Causeway Coast and Glens |
| **Lisburn** | LIS | 153 | 47377 | 21.61 | Lisburn and Castlereagh |
| **Lurgan** | LRG | 149 | 28634 | 16.01 | Armagh City, Banbridge and Craigavon |
| **Magherafelt** | MGF | 99 | 12017 | 8.49 | Mid Ulster |
| **Newry** | NEW | 152 | 34042 | 17.68 | Newry, Mourne and Down |
| **Newtownbreda** | NTB | 151 | 28693 | 14.49 | Lisburn and Castlereagh |
| **North Coast** | NCT | 139 | 42440 | 32.57 | Causeway Coast and Glens |
| **North Down** | NDN | 149 | 73384 | 27.91 | Ards and North Down |
| **Omagh** | OMA | 141 | 20200 | 16.31 | Fermanagh and Omagh |
| **Strabane** | STR | 136 | 13251 | 6.95 | Derry City and Strabane |
| **Warrenpoint** | WRP | 101 | 12766 | 6.65 | Newry, Mourne and Down |
| **Whitehouse** | WTH | 144 | 68952 | 28.54 | Antrim and Newtonabbey |

WW SARS-CoV-2 sample processing

*Sample collection*

Composite wastewater samples, comprising primary untreated influent were collected over a 24-hour period (November 14, 2021 to March 11, 2023) using an Isco Glacier autosampler (Isco; Lincoln, USA) from municipal WWTPs. These samples were provided by Northern Ireland Water Ltd and the Northern Ireland Environment Agency. The sampling timeline is depicted in Figure S1.

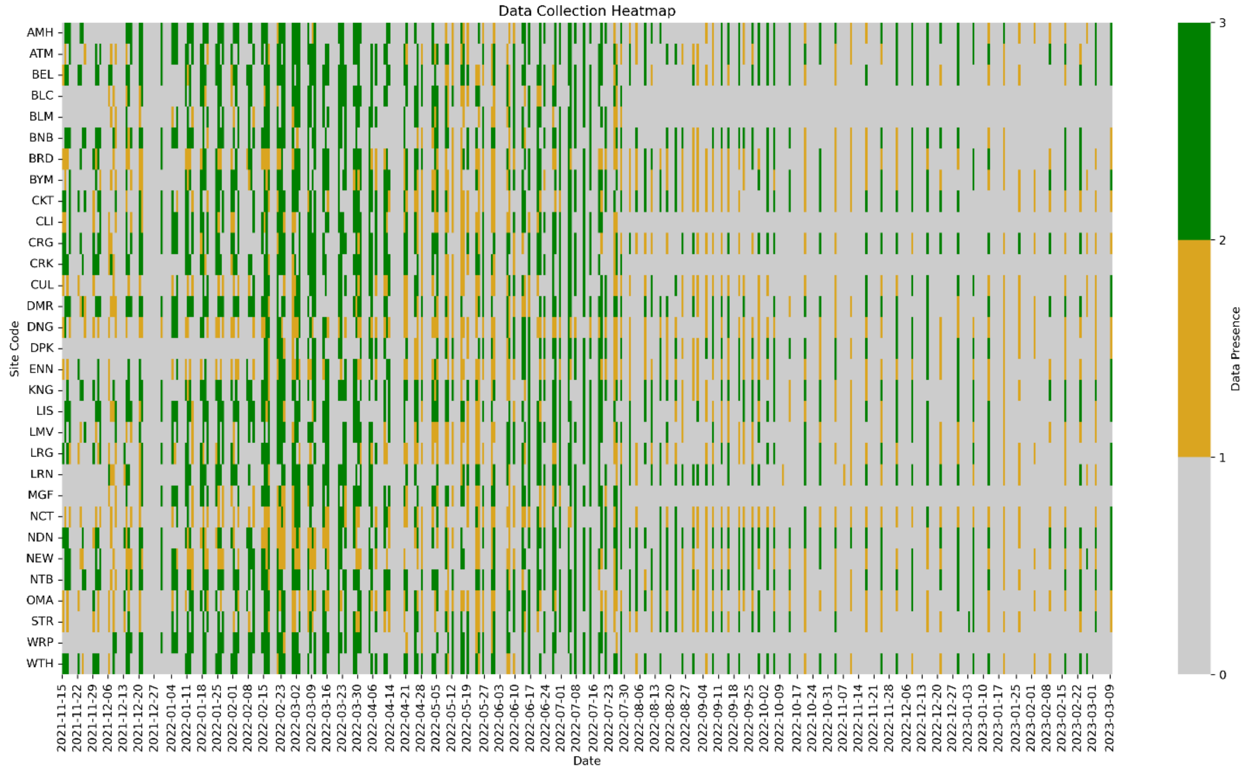

**Figure S1. Wastewater treatment plant (WWTP) sampling and quality control (QC) timeseries.** The y-axis represents each WWTP site, represented by its 3-letter short code, while the x-axis represents each date. Green indicates that a sample was collected and passed quality control (QC). Orange indicates that a sample was collected but failed QC by having less than 50% genome coverage. Gray indicates that no sample was collected.

*Sample processing and viral concentration*

Once received, primary influent wastewater samples were stored at 4 °C before pre-processing the same day. Wastewater (50mL) was clarified by centrifugation at 4,000 rpm for 10 mins (4°C). Wastewater supernatant was carefully transferred to a fresh 50mL tube before being concentrated using a CP-Select Concentrating Pipette™ using hollow fibre polysulfone high-flow pipette ultrafilter tips with a cut-off of 150 KDa (InnovaPrep LLC). Pipette tips were purged using an elution buffer containing 0.075% Tween 20 in 25 mM Tris buffer and the volume of concentrated eluates recorded. VetMAX™ Xeno™ Internal Positive Control (IPC) RNA (Applied Biosystems, ThermoFisher Scientific) was utilised as an internal control to monitor both nucleic acid recovery and PCR inhibition in purified wastewater samples.

*Nucleic acid extraction and purification*

Total nucleic acids were extracted and purified from 200 µL of concentrated wastewater sample, on a Roche MagNA Pure 96 Instrument using the DNA and Viral NA Small Volume Kit (Roche Diagnostic Limited) and the Pathogen Universal 200 extraction protocol. Purified nucleic acids were eluted in a 50 µL volume.

*SARS-CoV-2 RT-qPCR*

Extracted RNA was screened for the prescence of SARS-CoV-2 using AgPath-ID™ One-Step RT-qPCR Reagents (ThermoFisher Scientific) and SARS-CoV-2 N1 + N2 Assay Kits (Qiagen) in a final reaction volume of 25 µL. Each reaction included 12.5 µL 2x AgPath-ID™ RT-PCR Buffer, 1 µL 25x AgPath-ID™ RT-qPCR enzyme, 1 µL 20x primer/probe SARS-CoV-2 assay and 0.2 µL bovine serum albumin (0.2mg/mL, ThermoFisher Scientific). Four qPCR reactions (comprising 2x 10 µL and 2x 3 µL purified nucleic acid template volumes) were run per wastewater sample with amplification performed on a LightCycler 480 II Real-Time PCR System (Roche Diagnostic Limited) with the following thermo profile: 50°C for 10mins, 95°C for 10mins followed by 45 cycles of 95°C for 10s and 60°C for 30s. Positive and negative controls were included with each RT-PCR run, with all positive and negative controls, returning positive and negative results, respectively. Nucleic acid extracts were used as both template neat (2x 10 µL) and diluted (2x 3 µL), a widely applied strategy for the removal, or attenuation, of PCR inhibitors in stool samples[1]. All wastewater samples recorded as positive for SARS-CoV-2 were taken forward for sequencing.

*Whole-genome sequencing (WGS)*
Amplicon whole-genome sequencing of SARS-CoV-2 was performed following the Mini-XT SARS-CoV-2 protocol[2,3]. In brief, SARS-CoV-2 positive wastewater extractions and two negative controls (DEPC) were reverse transcribed using LunaScript® RT SuperMix Kit (New England Biolabs, Hitchin, UK) following the manufacturer's instructions. cDNA was amplified by tiled PCR (98 × 450 bp overlapping tiled amplicons, spanning the SARS-CoV-2 genome) using two primer pools from the ARTIC Network (ARTIC nCoV-2019 https://github.com/artic-network/primer-schemes/tree/master/nCoV-2019). The version of ARTIC primers was selected based on the changes in the dominant variants. cDNA amplification was done in two reactions (primer pool A and B) using Q5® Hot Start High-Fidelity DNA Polymerase (New England Biolabs). Following PCR, the amplicons from pools A and B were combined, purified using Kapa HyperPure beads (Roche Diagnostic Limited) and quantified using a Qubit fluorometer and dsDNA HS Assay Kit (ThermoFisher Scientific). The quantified product was normalised to a concentration of 0.2 ng/µl using the Echo 525 Liquid Handler (Beckman Coulter). Libraries were prepared using Nextera XT Library Preparation Kit with sequencing progressing on a MiSeq using v2 reagents and a 2 × 251 bp paired-end sequencing protocol (Illumina).

#### Individual sequencing programme

##### The healthcare landscape of Northern Ireland

Health and Social Care (HSC) is the publicly funded healthcare system in NI, which plays a central role in coordinating and delivering healthcare services across the region[4]. As the primary healthcare authority, HSC has been responsible for managing various aspects of the COVID-19 response, including population testing and genomic surveillance initiatives.

Under the umbrella of HSC, there are six Health and Social Care Trusts (HSCTs) operating in NI. These trusts are responsible for delivering health and social care services to specific geographic areas within the region. Among these HSCTs, five are regional trusts, each serving a distinct area, while the ambulance trust provides emergency medical services across the entirety of NI. For futher details about each HSCT, please refer to Table S2 and Figure S2 below.

The HSC system in Northern Ireland comprises various entities responsible for delivering healthcare services. Among these entities are the Health & Social Care Trusts (HSCTs), which are administrative bodies overseeing specific geographic areas within the HSC system. While HSC represents the broader healthcare framework, HSCTs are individual trusts responsible for managing and delivering health and social care services at a local level.

**Table S2. Health & Social Care Trust (HSCT) Information.**
This table summarizes key information about each Health & Social Care Trust (HSCT) in Northern Ireland, including the population served, regional area coverage, number of staff, and number of healthcare facilities.

| **Health & Social Care Trust (HSCT)** | **Abbreviation** | **Population Served (2011 census)** | **Regions Served** | **Regional Area (EPSG: 2157) [km$^2$]** | **Number of staff (estimated)** |
|---|---|---|---|---|---|
| Belfast HSCT | BHSCT | 348,204 | Belfast, Castlereagh | 200 | 22,000 |
| Northern HSCT | NHSCT | 463,297 | Antrim, Ballymena, Ballymoney, Carrickfergus, Coleraine, Cookstown, Larne, Magherafelt, Moyle, Newtownabbey | 4,380 | 12,000 |
| South Eastern HSCT | SEHSCT | 346,911 | Ards, Down, North Down, Lisburn | 4,839 | 10,000 |
| Southern HSCT | SHSCT | 380,312 | Armagh, Banbridge, Craigavon, Dungannon, Newry and Mourne | 3,186 | 13,000 |
| Western HSCT | BHSCT | 294,417 | Derry/Londonderry, Fermanagh, Limavady, Omagh, Strabane | 1,703 | 12,000 |
| Northern Ireland Ambulance Service HSCT | NIAS | 1,833,141 | All | 14,308 | 1,300 |

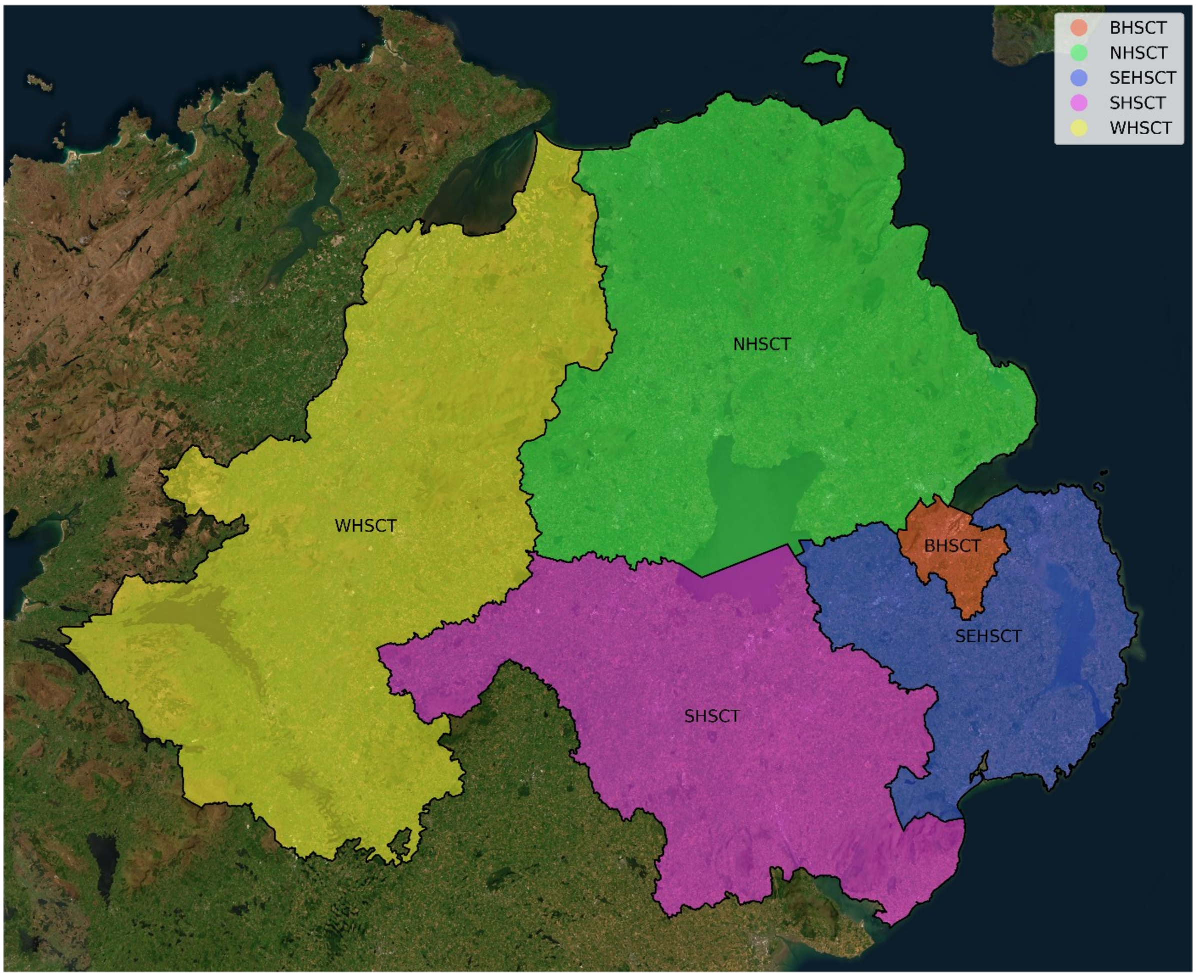

**Figure S2. Map of Northern Ireland's five regional Health & Social Care Trusts (HSCTs)**
This map illustrates the geographical distribution of the five regional HSCTs across Northern Ireland. The geospatial vector data to depict this map was obtained from OpenDataNI (https://admin.opendatani.gov.uk/dataset/department-of-health-trust-boundaries).

#### COG-UK collaboration

Researchers in NI actively participated in the COVID-19 Genomics UK Consortium (COG-UK), a collaborative effort aimed at sequencing SARS-CoV-2 genomes across the United Kingdom from April 2020 to March 2023[5]. COG-UK collected samples from various sources, integrated genome data with epidemiological information, and conducted research to monitor virus spread, assess changes in transmissibility and virulence, and evaluate treatment effectiveness. It transitioned to a public health-led national service from April to September 2021, focusing thereafter on data linkage, research, and international training. The pioneering work of COG-UK in large-scale sequencing and data sharing had a significant impact on global efforts during the pandemic's early years, including in NI. Protocols and tools developed by COG-UK consortium members were instrumental in enhancing NI's genomic surveillance capabilities, contributing to pandemic preparedness and ongoing research efforts within the region.

#### Individual testing

Nasopharyngeal swabs from individuals undergo RT-qPCR testing, and positive samples are forwarded for whole-genome sequencing (WGS) as part of the UK Department of Health and Social Care's multifaceted approach. Pillar 1 testing, centred on HSC labs and hospitals, focuses on patients and frontline workers, while Pillar 2 extends testing to the wider population for effective COVID-19 management. Pillar 2 sampling in NI has ceased during June 2022.

#### Whole-genome sequencing (WGS)

The dataset of genomes studied focuses on analysing 22,924 SARS-CoV-2 genomes sequenced with Illumina and 556 with Nanopore[6], all with complete metadata, which were collected between November 14, 2021 and March 11, 2023.

We utilised the Mini-XT protocol for the vast majority of the sequences presented here[2,3]. In brief, SARS-CoV-2 viral nucleic acid extracts from Pillar 1 and Pillar 2 sources were converted to complementary DNA (cDNA) and subsequently, the cDNA was amplified by tiled PCR using separate primer pools from the ARTIC primer panels (i.e., V4, V4.1, V5.2, V5.3.2)[7,8]. The resulting amplicons were combined, purified by bead cleaning, and then eluted and quantified with Quant-iT™ dsDNA broad range kit (Invitrogen™, ThermoFisher Scientific). These quantified PCR products were concentration-normalised to 0.2 ng/µL using the Echo Liquid Handler and the libraries were prepared using Nextera XT Library Preparation Kit (Illumina Ltd., Cambridge, UK). The library pool was bead cleaned up and then diluted, denatured, and loaded onto the Illumina MiSeq system for 151 bp paired-end sequencing. FASTQ generation was conducted onboard, with secondary bioinformatic processing occurring offboard (see ***Bioinformatics*** below).

For the subset of samples sequenced using Oxford Nanopore technology, 50ng of purified PCR product for each sample was end-prepared and indexed using the SQK-LSK109 ligation kit (Oxford Nanopore Technologies, UK) in combination with native barcoding expansion kits SQK-NBD104/SQK-NBD114 (Oxford Nanopore Technologies, UK). Pooled libraries were cleaned using KAPA Pure magnetic beads (Roche, UK), and sequencing adapter ligation performed in accordance with manufacturer's instructions. Final libraries were loaded onto R9.4.1 MinION flow cells (Oxford Nanopore Technologies, UK) and sequenced on the GridION platform.

#### Geographic Information Systems (GIS)

##### Geospatial mapping

Geopandas v0.14.3[9] was used to generate geographic visualizations within the manuscript and to support geospatial filtering and analyses. The Esri "World Imagery" basemap (Sources: Esri, DigitalGlobe, GeoEye, i-cubed, USDA FSA, USGS, AEX, Getmapping, Aerogrid, IGN, IGP, swisstopo, and the GIS User Community) as retrieved using contextily v1.6.0 (https://github.com/geopandas/contextily) is also used to provide satellite-based imagery of the region.

##### Local government districts (LGDs) in Northern Ireland

Eleven local government districts (LGDs) constitute NI. These LGDs are unitary administrations responsible for all aspects of local government in NI. For our geospatial analysis at the LGD-level, we obtained a shapefile from OpenDataNI (https://admin.opendatani.gov.uk/dataset/osni-open-data-largescale-boundaries-local-government-districts-2012) to map the 11 LGDs (LGD2014), which were delineated in 2012.

**Table S3. LGD Abbreviations and Associated HSC Trust(s)**

| Local government district (LGD) | Troendle-Rice-Simpson-Skvortsov abbreviation[10] | HSC Trust(s) | WWTPs sampled |
|---|---|---|---|
| **Antrim and Newtownabbey** | AN | Northern | 3 |
| **Ards and North Down** | AD | South Eastern | 3 |
| **Armagh City, Banbridge and Craigavon** | AB | Southern | 4 |
| **Belfast** | BF | Belfast | 1 |
| **Causeway Coast and Glens** | CC | Northern, Western | 3 |
| **Derry City and Strabane** | DS | Western | 2 |
| **Fermanagh and Omagh** | FO | Western | 2 |
| **Lisburn and Castlereagh** | LC | South Eastern, Belfast | 3 |
| **Mid and East Antrim** | AM | Northern | 3 |
| **Mid Ulster** | MU | Northern, Southern | 4 |
| **Newry, Mourne and Down** | NM | Southern, South Eastern | 3 |

#### Bioinformatics

##### Nextflow pipeline (ncov2019-artic-nf)

Bioinformatics analyses were conducted using the Illumina Nextflow pipeline[11] developed by the ARTIC network, tailored for the processing of SARS-CoV-2 sequencing data. The pipeline automates the execution of the ARTIC network's fieldbioinformatics tools (https://github.com/artic-network/fieldbioinformatics), designed for the analysis of viral genomic data, as routinely utilised by members of COG-UK. The version utilised for this study is available in the forked GitHub repository at https://github.com/QUB-Simpson-lab/ncov2019-artic-nf.

*Software updates and inclusions*

To enhance functionality of the bioinformatics analysis, we made several upgrades and inclusions compared to the original Nextflow pipeline version (https://github.com/connor-lab/ncov2019-artic-nf):

**Software Version Upgrades:**

- SAMtools[12] (https://github.com/samtools/samtools) & BCFtools[13] (https://github.com/samtools/bcftools) were upgraded from version 1.10 to 1.18.
- trim_galore (https://github.com/FelixKrueger/TrimGalore) was upgraded from version 0.6.5 to 0.6.10.
- iVar[14] (https://github.com/andersen-lab/ivar) was upgraded from version 1.3 to 1.4.2.

**Inclusion of Freyja and Pangolin:**

- Freyja[15] version 1.4.9 (https://github.com/andersen-lab/Freyja), for depth-weighted demixing and variant calling, and Pangolin[16] version 4.3.1 (https://github.com/cov-lineages/pangolin) (pangolin-data version 1.25.1), for lineage calling, were integrated directly into the pipeline.

*Pipeline workflow*

Within the Nextflow pipeline, the following steps were executed:

1. Reference Genome Preparation:
    - The SARS-CoV-2 reference genome sequence[17] FASTA (MN908947.3) and ARTIC primer scheme[8] Browser Extensible Data[18] (BED) files (https://github.com/artic-network/primer-schemes) were downloaded.
    - The reference genome was indexed using the BWA[19] (v0.7.17) index tool.
2. Pre-processing of FASTQ Files:
    - For each pair of paired-end (R1/R2) FASTQ files, rim_galore, a wrapper around Cutadapt[20] and FastQC[21], was employed to ensure consistent adapter and quality trimming of FASTQ files.
3. Read Mapping:
    - BWA[19] mem was used to map the adapter and quality trimmed reads to the reference genome.
4. Primer Sequence Removal:
    - iVar[14] trim was applied with the ARTIC primer scheme BED file to remove any mapped primer sequences from the aligned reads.
5. Consensus Sequence Generation:
    - Consensus sequences were generated using iVar[14] consensus by passing the output of samtools mpileup for the BAM file.
6. Variant Calling:
    - Variants were called using iVar[14] variants, leveraging the output of SAMtools[12] mpileup for the BAM file.
    - Additionally, Freyja[15] variants is used to create an unfiltered variants file along with a depth file.
7. Depth-Weighted Demixing with Freyja:
    - Freyja[15] demix was utilised within the pipeline for depth-weighted demixing. The --depthcutoff parameter in Freyja was configured to 10 to mitigate demixing errors that may arise during Freyja's resolution of the constrained (unit sum, non-negative) demixing problem using the Embedded Conic Solver (ECOS)[22] of CVXPY[23,24].
8. Lineage Calling with Pangolin:
    - Pangolin[16] was employed to call the Pango lineage[25] of the consensus sequence obtained upstream.

A flowchart of the pipeline can be found on the following page as Figure S3.

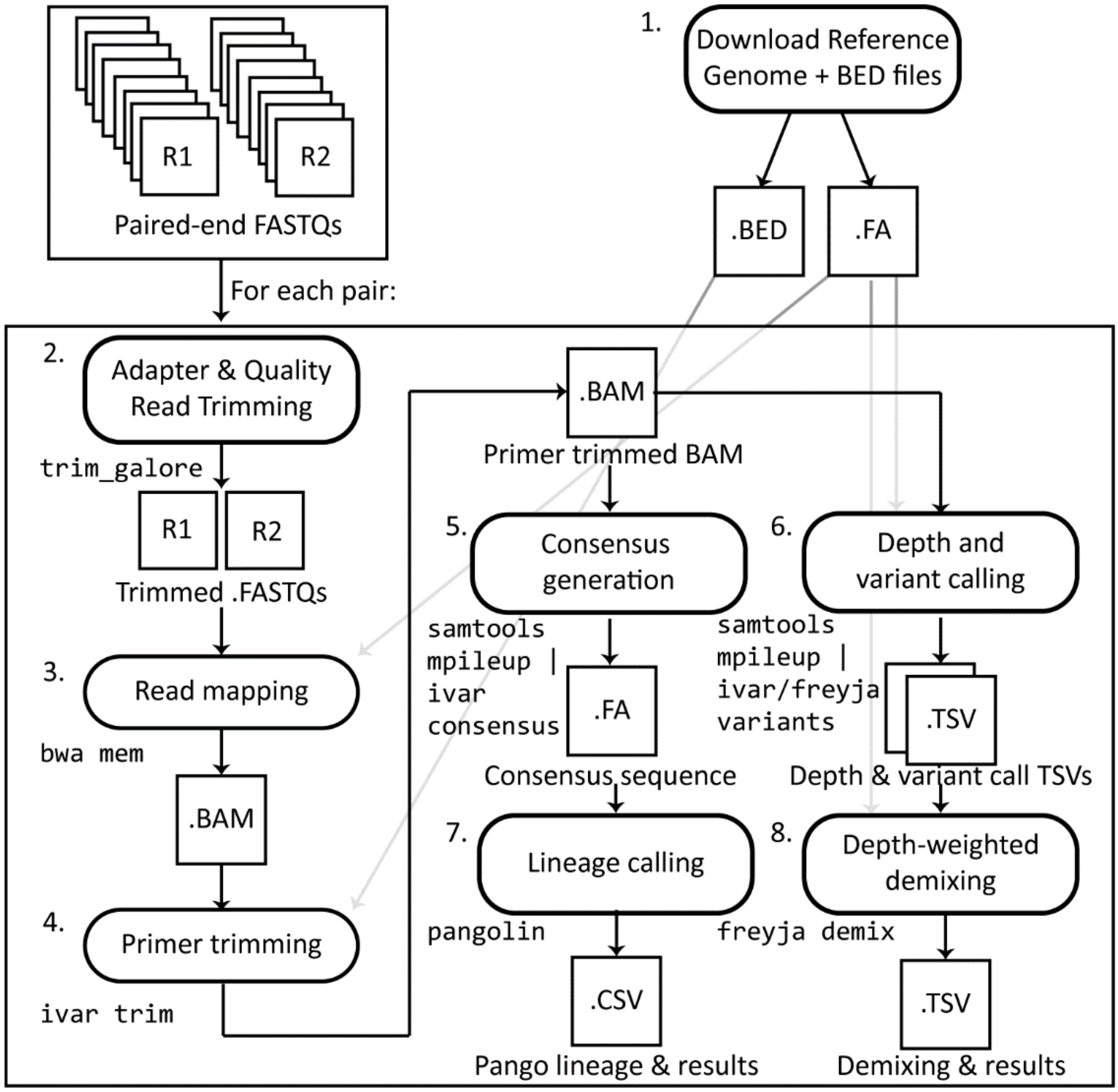

**Figure S3. ncov2019-artic-nf pipeline data flow chart.**

#### Classification of SARS-CoV-2 variants into mutation constellations using regular expression patterns

To classify SARS-CoV-2 variants into mutation constellations , we utilised regular expression patterns[26] to parse and categorize Pango[25] lineage names. Constellations represent collections of mutations with functional significance that may arise independently multiple times within the virus's genome[27]. This classification organized the Pango lineages as demixed by Freyja[15] and identified by pangolin[16], as outlined in Table S4 below.

**Table S4. SARS-CoV-2 mutation constellation designations**

<table>
<tr><th>Mutation constellation moniker</th><th>Included Pango lineages</th><th>Regex pattern</th></tr>
<tr><td>Alpha-like</td><td>B.* (excl. B.1.1.529 & B.1.617) /AZ.*/V.*/Q.*</td><td>^(B\.(?!1\.1\.529|1\.617)|AZ\.|C\.|P\.|Q\.|V\.).*</td></tr>
<tr><td>Delta-like</td><td>B.1.617*/AY*</td><td>^((AY)|(B\.1\.617)).*</td></tr>
<tr><td>Omicron BA.1-like</td><td>BA.1*/BC.*/BD.*</td><td>B((A\.1)|(\.1\.1\.529)|[CD]\.).*</td></tr>
<tr><td>Omicron BA.2-like</td><td>BA.2* (excl. BA.2.75*)/BG.*/DD.*/DS.*</td><td>^(BA\.2(?!\.75)|BG\.|D[DS]\.).*</td></tr>
<tr><td>Omicron BA.2.75-like</td><td>BA.2.75*/BL.*/BM.*/BN.*/BR.*/BY.*/CA.*/CB.*/CH.*/CJ.*/CV.*/DE.*/DV.*/EJ.*/GP.*</td><td>^(BA\.2\.75|B[LMNRY]\.|C[ABHJV]\.|D[EV]\.|EJ\.|GP\.).*</td></tr>
<tr><td>Omicron BA.3-like</td><td>BA.3*</td><td>^BA\.3.*</td></tr>
<tr><td>Omicron BA.4-like</td><td>BA.4*</td><td>^BA\.4.*</td></tr>
<tr><td>Omicron BA.5-like</td><td>BA.5*/BE.*/BF.*/BK.*/BT.*/BV.*/BZ.*/CE.*/CF.*/CG.*/CK.*/CP.*/CQ.*/CR.*/CT.*/CU.*/CW.*/DA.*.DH.*/DL.*/DP.*/DT.*/DU.*/DZ.*/EB.*/ED.*/EE.*/EF.*/EN.*/EQ.*/ER.*/EW.*/FM.*</td><td>^((B(A\.5|[EFKTVZ]\.))|C[EFGKPQRTUW]\.|D[AHLPTUZ]\.|E[BDEFNQRW]\.|FM\.).*</td></tr>
<tr><td>Omicron BQ-like</td><td>BQ*</td><td>^BQ\..*</td></tr>
<tr><td>Recombinants</td><td>X* (not XBB)</td><td>^X(?!BB).*</td></tr>
<tr><td>Omicron XBB-like</td><td>XBB*/EG*/EK*/EU*/FE*/FL*/FY*/GA*/GF*/GK*/JY*/HK*/HT*/HV*/JD*/JG*/JK*</td><td>^((XBB)|(E[GKU])|(F[ELY])|(G[AFKY])|(H[KTV])|(J[DGK])).*</td></tr>
<tr><td>Other</td><td>All others observed</td><td></td></tr>
</table>

#### Geospatial abundance timeseries calculations of SARS-CoV-2 constellations

We implemented a computational metadata-based sample extraction and analysis approach using pandas v2.2.2[28] to estimate the abundance of SARS-CoV-2 constellations, which is crucial for understanding viral dynamics and epidemiological trends across various regions of interest, such as all of Northern Ireland, specific Local Government Districts (LGDs) or wastewater treatment plant (WWTP) catchment areas. We retrieved all geographically relevant samples within specified time ranges by accessing sample metadata from the Cloud Infrastructure for Microbial Bioinformatics (CLIMB) project. Our active participation in the project as part of our involvement in COG-UK included sequencing the data and uploading it to the CLIMB platform. Additionally, individual-level metadata provided by Northern Ireland's HSC Public Health Agency (PHA) was mirrored to GISAID. This metadata, combined with our sequencing efforts, facilitated a comprehensive analysis of SARS-CoV-2 genomes and their anonymised epidemiological context. Each individual sample was mapped to a constellation from its consensus Pango[25] lineage, or the set of constellations and summed abundances for the wastewater samples as demixed by Freyja[15]. Subsequently, we computed either daily counts for individual sequences or daily means for wastewater samples for each constellation category, ensuring a continuous time series through linear interpolation. To mitigate data noise and fluctuations, we applied a centred uniform rolling window smoothing of 15 days (± 1 week). Percentage normalization was then applied to facilitate comparisons across different dates, regions, and sequencing programmes. Finally, we visualized the aggregated abundances and their smoothed percentages over time using Matplotlib v3.8.4[29] and Seaborn v0.13.2[30], which aided in the interpretation of geospatiotemporal abundance trends and patterns. This approach could also be applied to track the dynamics of individual variants and mutations.

#### Optimal temporal alignment analysis for comparative assessment of SARS-CoV-2 constellation abundances

To determine lag and leads across entire SARS-CoV-2 constellation abundance timeseries between regions or programmes, we developed an optimal temporal alignment analysis algorithm. This algorithm compares pairs of time series for a given SARS-CoV-2 constellation as obtained from either different regions within a chosen sequencing programme or between different sequencing programmes within the same region. Utilising a systematic shifting approach, we assess potential lags or leads by shifting one time series relative to the other across a range of possible daily shifts (e.g., ± 30 days). Calculating the mean absolute error at each shift quantifies the discrepancy between them, and through iterating over all possible shifts, the optimal alignment is identified by evaluating the minimum. See the Supporting Information for a formal mathematical description of the mean absolute error as well as pseudocode for the shifting algorithm.

#### Utilising the Freyja UShER substitution barcode database for detailed set-based nucleotide substitution tracking during constellation transitions within wastewater

Set-based nucleotide substitution tracking over constellation transitions is informed by the Freyja UShER substitution barcode database, a repository of genomic substitutions within SARS-CoV-2 genomes included with and produceable by the software (Freyja update). Organized in a matrix format, each column corresponds to a specific genomic substitution (e.g., G210T), denoted by REF-POS-ALT, where REF is the reference nucleotide, POS is the 1-based position of the substitution within the genome, and ALT is the altered nucleotide. Each row in the matrix represents a unique Pango lineage of SARS-CoV-2, with presence or absence of a substitution indicated by binary values: 0 for absence and 1 for presence.

For constellation transitions, the process begins by constructing comprehensive sets of substitutions for each constellation. These sets are derived by filtering rows from the Freya UShER substitution barcode database file based on regex patterns corresponding to the constellation's lineages (as outlined in Table S3), and then selecting all columns (substitutions) with at least one non-zero value, ensuring the inclusion of all substitutions considered by Freyja to define the lineages that it finds through demixing that contribute the constellation. The sets are further refined by retaining only substitutions present in all lineages of the constellation (i.e., all rows are 1), providing a more conservative depiction of the constellation's genetic profile.

Derived from the comprehensive and refined sets of substitutions for each constellation involved in a given transition, eight distinct subsets are created to comprehensively capture diverse mutation dynamics across constellation transitions:

1. Substitutions unique to all lineages in the initial constellation.
2. Substitutions unique to all lineages in the subsequent constellation.
3. Substitutions unique to some lineages in the initial constellation.
4. Substitutions unique to some lineages in the subsequent constellation.
5. Substitutions present in all lineages in the initial constellation and some in the subsequent constellation.
6. Substitutions present in all lineages in the subsequent constellation and some in the initial constellation.
7. Substitutions present in all lineages in both constellations.
8. Remaining substitutions present in some lineages of both constellations.

The allele frequencies for every substitution in each subset are then monitored over time, utilising data extracted from the wastewater samples' iVar variants and depth of sequencing TSV (Tab-Separated Values) files as generated by the Freyja variants submodule of the bioinformatic pipeline. The files provide detailed information on the sequencing depth of aligned and trimmed reads, as well as the detected variant alleles in the samples, including their alternate allele frequencies. Substitutions are considered for analysis only if they have a minimum read depth of 10 at the specific nucleotide position in the genome. If the read depth falls below this threshold, the allele frequency data for that substitution in the corresponding sample is disregarded. Subsequently, the allele frequency data timeseries data are processed by averaging, interpolating, and smoothing the readouts using methods consistent with those employed for the constellation abundance time series described earlier. This data processing ensures a coherent and standardized analysis approach across all datasets.

This approach facilitates a comprehensive analysis of mutation patterns and evolutionary dynamics of SARS-CoV-2 as utilised by Freyja across constellation transitions. For a visual representation of the set-based approach, please refer to Figure S4A. Refer to the Supporting Information for a comprehensive delineation of the substitution sets tracked in each transition.

#### Customisation of Freyja UShER barcodes

During transitions between major constellation lineage groups (e.g., Delta-like to Omicron BA.1-like), we observed instances where Freyja[15] initially reported a high abundance of recombinant lineages in wastewater samples. This ambiguity arose from Freyja's challenge in distinguishing between mixed samples of parent lineages and their associated recombinants. To address this, we produced a customized Freyja UShER[31] substitution barcodes file that excludes recombinant Pango[25] lineages (X*) while retaining XBB* lineages, which were observed as a dominant constellation within the individual sequences towards the end of the study. By instructing Freyja to exclude other recombinant lineages during demixing, we successfully salvaged abundances into the relevant major lineage families (constellations).

#### ADDITIONAL SUPPORTING INFORMATION:

##### Effects and considerations of excluding sets of recombinant lineages from Freyja's demixing of wastewater samples

Figure S3 provides a visual representation of the effects of excluding recombinant lineages from Freyja's analysis by comparing constellation abundance timeseries before and after customization. While this customization has enhanced the accuracy of lineage identification in our study, certain limitations must be acknowledged. One such limitation is the potential exclusion of rare recombinant lineages that could offer insights into viral evolution and transmission dynamics. Focusing solely on predominant parent lineages may overlook rare recombinants crucial for understanding emergent variants and genetic dissemination.

Freyja's detection of recombinants in a mix of parent lineages can lead to false positives due to inherent mathematical challenges. These challenges encompass both sequencing noise and the complexities of distinguishing between mixtures of recombinants and parent lineages.

Sequencing noise can exacerbate the difficulty of accurately identifying sublineages, particularly when dealing with subtle sequence variations. Additionally, the presence of particular single nucleotide polymorphisms (SNPs) within the UShER barcodes can further complicate matters. As many SNPs may belong to multiple lineages, they appear in both recombinant and parent lineages. This ambiguity can often make it challenging for Freyja to determine whether or to what degree a sample represents a mix of parent lineages or a combination of recombinants and parent lineages. For instance, identifying both recombinants and their parent lineages alongside each other could pose challenges, as the presence of shared SNPs between recombinants and parent lineages may confound lineage assignment.

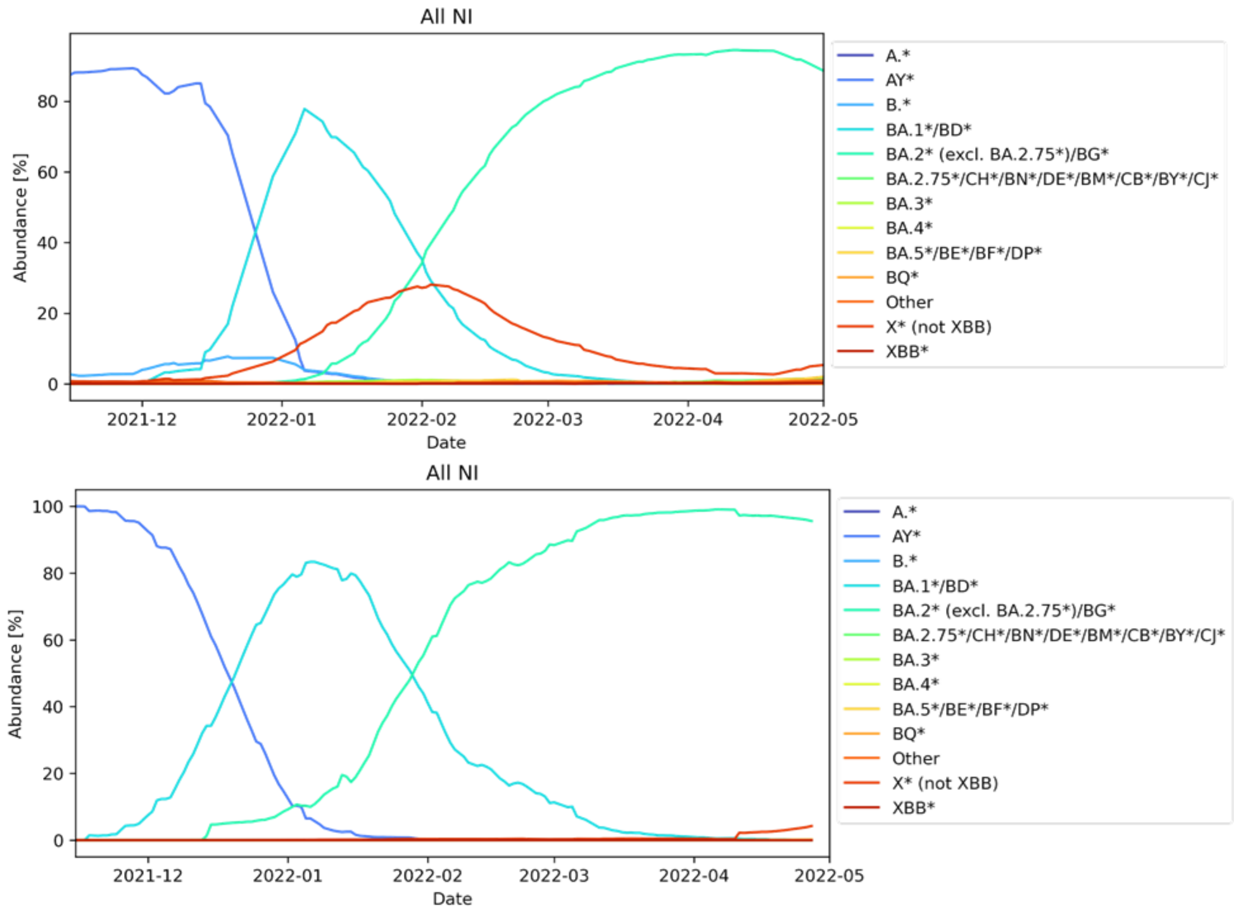

**Figure S4. Effect of removing X* lineages from Freyja detections in the WW between 14th Nov 2021 and 1st May 2022.**

#### Calculation of the mean absolute error (MAE)

Consider two time series denoted as $X = \{x_1, \dots, x_n\}$ and $Y = \{y_1, \dots, y_n\}$, each containing $n$ data points. In these time series, each $x_i$ and $y_i$ represents the intensity of the signal at the $i^{\text{th}}$ sampled time. The mean absolute error ($MAE$) between timeseries $X$and $Y$ is:

$$MAE(X,Y) = \frac{1}{n}\sum_{i=1}^{n}|y_i - x_i|$$

Here,

- $|\cdot|$ denotes the absolute value or modulus operation, representing the positive distance between the two values.
- The summation $\sum_{i=1}^{n}$ is performed over each of the corresponding $n$ datapoints in succession.
- The prefactor $\frac{1}{n}$ provides the mean or average of the summation result.

**Error! Reference source not found.** quantifies the average absolute difference between corresponding data points of the two time series, providing a measure of their dissimilarity.

#### Pseudocode for the optimal temporal alignment algorithm

In many scientific and engineering applications, comparing two curves or datasets is essential for identifying similarities or discrepancies between them. One commonly used method for this purpose is to find the shift value that minimizes the deviation between the curves. This shift value indicates the optimal alignment between the curves, providing the best estimate of the temporal displacement between them. Determining this optimal alignment is crucial for accurately assessing the temporal relationship between the curves and quantifying any temporal discrepancies. It ensures that the second curve is correctly positioned relative to the first, facilitating precise analysis of their temporal dynamics

Here, we present an algorithmic approach used in our study to find the best shift value between two curves. The algorithm systematically evaluates different shift values and selects the one that results in the lowest mean absolute error. This approach ensures that the curves are optimally aligned, facilitating accurate comparison and analysis.

The pseudocode provided below outlines the steps of the algorithm, along with supporting functions for padding arrays, calculating the mean absolute error, and computing the mean of arrays with NaN (Not a Number) values. Padding with NaN values is essential to ensure that the shifting process is not influenced by the introduction of additional data points. By using NaN values for padding, we isolate the shifted sections of the curves from the original data, thereby preventing any distortion in the calculation of the mean absolute error. This practice ensures accurate alignment between curves and enhances the integrity of the comparative analysis.

```
function find_best_shift_value(y0, y1, max_shift):
    best_shift = 0
    min_error = infinity
    for shift_val from -max_shift to max_shift:
        error = mean_absolute_error(y0, y1, shift_val)
        if error < min_error:
            min_error = error
            best_shift = shift_val
    return best_shift
function mean_absolute_error(y0, y1, shift_val):
    if shift_val > 0:
        new_y0 = pad_end(y0, shift_val, nan)
        new_y1 = pad_begin(y1, shift_val, nan)
    else if shift_val < 0:
        pos_shift = abs(shift_val)
        new_y0 = pad_begin(y0, pos_shift, nan)
        new_y1 = pad_end(y1, pos_shift, nan)
    else:
        return nanmean(abs(y1 - y0))
    return nanmean(abs(new_y1 - new_y0))
function pad_end(arr, num, val):
    N = length(arr)
    new_arr = create_array(N + num)
    copy(arr, new_arr)
    fill_end(new_arr, num, val)
    return new_arr
function pad_begin(arr, num, val):
    N = length(arr)
    new_arr = create_array(N + num)
    fill_begin(new_arr, num, val)
    copy(arr, new_arr[num:])
    return new_arr
function fill_end(arr, num, val):
    for i from length(arr) - num to length(arr) - 1:
        arr[i] = val
function fill_begin(arr, num, val):
    for i from 0 to num - 1:
        arr[i] = val
function copy(src, dest):
    for i from 0 to length(src) - 1:
        dest[i] = src[i]
function create_array(size):
    return array of size elements
function length(arr):
    return number of elements in arr
function nanmean(arr):
    sum = 0
    nan_count = 0
    for each element in arr:
        if isnan(element):
            nan_count = nan_count + 1
        else:
            sum = sum + element
    return sum / (length(arr) - nan_count)
```

**Table S5. Complementarity of individual and WW WGS (see Figure 4).**

| Region | Period | Core WW | Core Individuals (Indiv.) | Core Both | Accessory (Acc.) WW | Acc. Indiv. | Acc. Both | Other WW | Other Indiv. | Other Both |
|---|---|---|---|---|---|---|---|---|---|---|

| | | | | | | | | | | |
|---|---|---|---|---|---|---|---|---|---|---|
| **AB** | <01May22 | 2 | 4 | 85 | 54 | 603 | 203 | 803 | 1814 | 292 |
| | ≥01May22 | 26 | 3 | 70 | 283 | 20 | 33 | 1053 | 52 | 17 |
| **AD** | <01May22 | 0 | 8 | 82 | 55 | 533 | 157 | 776 | 1500 | 221 |
| | ≥01May22 | 1 | 4 | 96 | 175 | 98 | 85 | 851 | 237 | 53 |
| **AM** | <01May22 | 1 | 5 | 82 | 40 | 508 | 126 | 391 | 1435 | 158 |
| | ≥01May22 | 2 | 6 | 95 | 74 | 173 | 91 | 420 | 312 | 52 |
| **AN** | <01May22 | 0 | 8 | 84 | 26 | 578 | 146 | 321 | 1725 | 175 |
| | ≥01May22 | 3 | 4 | 97 | 129 | 145 | 102 | 532 | 289 | 62 |
| **BF** | <01May22 | 0 | 14 | 82 | 13 | 857 | 105 | 220 | 2818 | 126 |
| | ≥01May22 | 3 | 6 | 95 | 49 | 201 | 84 | 256 | 366 | 39 |
| **CC** | <01May22 | 1 | 7 | 82 | 46 | 504 | 142 | 600 | 1440 | 155 |
| | ≥01May22 | 1 | 6 | 97 | 114 | 110 | 71 | 548 | 281 | 43 |
| **DS** | <01May22 | 0 | 10 | 79 | 24 | 603 | 66 | 296 | 1595 | 71 |
| | ≥01May22 | 27 | 2 | 70 | 114 | 38 | 18 | 322 | 41 | 9 |
| **FO** | <01May22 | 1 | 6 | 81 | 24 | 412 | 85 | 317 | 1157 | 79 |
| | ≥01May22 | 11 | 3 | 83 | 84 | 91 | 40 | 329 | 129 | 17 |
| **LC** | <01May22 | 0 | 9 | 83 | 44 | 516 | 181 | 494 | 1640 | 210 |
| | ≥01May22 | 16 | 1 | 87 | 198 | 95 | 87 | 717 | 225 | 64 |
| **MU** | <01May22 | 1 | 11 | 83 | 57 | 500 | 157 | 676 | 1540 | 157 |
| | ≥01May22 | 2 | 15 | 84 | 103 | 149 | 80 | 466 | 236 | 41 |
| **NM** | <01May22 | 0 | 4 | 82 | 30 | 646 | 109 | 475 | 1894 | 141 |
| | ≥01May22 | 3 | 4 | 95 | 132 | 101 | 83 | 588 | 214 | 47 |
| **NI** | <01May22 | 1 | 10 | 91 | 81 | 824 | 752 | 3566 | 6568 | 1830 |
| | ≥01May22 | 4 | 0 | 107 | 378 | 230 | 513 | 4393 | 1018 | 682 |

#### Mathematical description of detecting transitions in variant constellation compositions over time

Let $C_i$ denote the composition vector of the constellations on day $i$, where $i = 1,2, \ldots, N$ and $N$ represents the total number of days in the period. Each composition vector $C_i$ is an $m$-dimensional vector, where $m$ is the total number of constellations present.

To quantify the change in constellation composition from day $i$ to day $i+1$, we compute the absolute difference between the composition vectors of adjacent days:
$\Delta C_i = |C_{i+1} - C_i|$, where $|\cdot|$ denotes element-wise absolute difference.
The overall change in variant composition for each day $i$ is then calculated by summing the absolute differences across all variants:

$$\boldsymbol{Net\ Overall\ Change_i} = \sum_{j=1}^{m} \Delta \boldsymbol{C_{i,j}}$$

, where $\Delta C_{i,j}$ represents the $j^{\text{th}}$ element of $\Delta C_i$
Repeating this process daily in a series of measurements for $Net\ Overall\ Change$, with representing the overall change in variant composition between each successive date in the period.
By setting a threshold $T$ on the values of $Net\ Overall\ Change$,, transitions in variant composition can be identified. For days ($i$) when $Net\ Overall\ Change_i \geq T$, it indicates a period of reckonable change in the mix of constellations, signifying a transition. Conversely, values of $Net\ Overall\ Change_i$ below $T$ signify more stable periods.

#### Wastewater surveillance shows both early detection of substitutions associated with emerging constellations and prolonged presence of substitutions from fading constellations

In our analysis, we observed earlier detection of substitutions belonging to rising constellations and extended shedding of substitutions belonging to disappearing constellations in wastewater sample by tracking allele frequencies in comparison to that obtained by abundance predictions from Freyja demixing. This phenomenon, characterized by prolonged presence and elevated levels of nucleotide substitutions in wastewater, poses challenges to traditional demixing approaches and affects the accuracy of tracking incoming variants.

To investigate these dynamics, we utilised a set-based nucleotide substitution tracking method employing the Freyja UShER substitution barcode database. This approach allowed detailed tracking of substitutions across constellation transitions, providing insights into evolutionary patterns within SARS-CoV-2. For each substitution subset, allele frequencies were monitored over time using data extracted from wastewater samples' iVar variants and depth TSV files generated by the Freyja bioinformatic pipeline (Freyja variants). Allele frequency time series data were processed identically as for the constellation abundance time series to ensure coherence and standardization, facilitating a detailed analysis of mutation patterns and evolutionary dynamics across constellation transitions.

Figure S4A illustrates the set-based substitution tracking scheme employed in our analysis. Substitutions were categorized based on their presence across constellations, providing insights into their dynamics during transitions. The figure also presents allele frequency dynamics for select transitions, highlighting the mean frequencies of each subset over time. In each of the transitions observed (Figure S4B-G), we note that the blue substitution set shows a shift in allele frequency relatively leftwards (i.e., earlier in time), while the red substitution set remains at higher frequency beyond that expected from the mean of substitutions as demixed from Freyja. This highlights, albeit with retrospective insight, the potential of wastewater surveillance to detect specific substitutions belonging to the incoming constellation earlier (blue), while also facilitating the assessment of the temporal dynamics of viral shedding, focusing on substitutions associated with the previously dominant constellation (red).
.

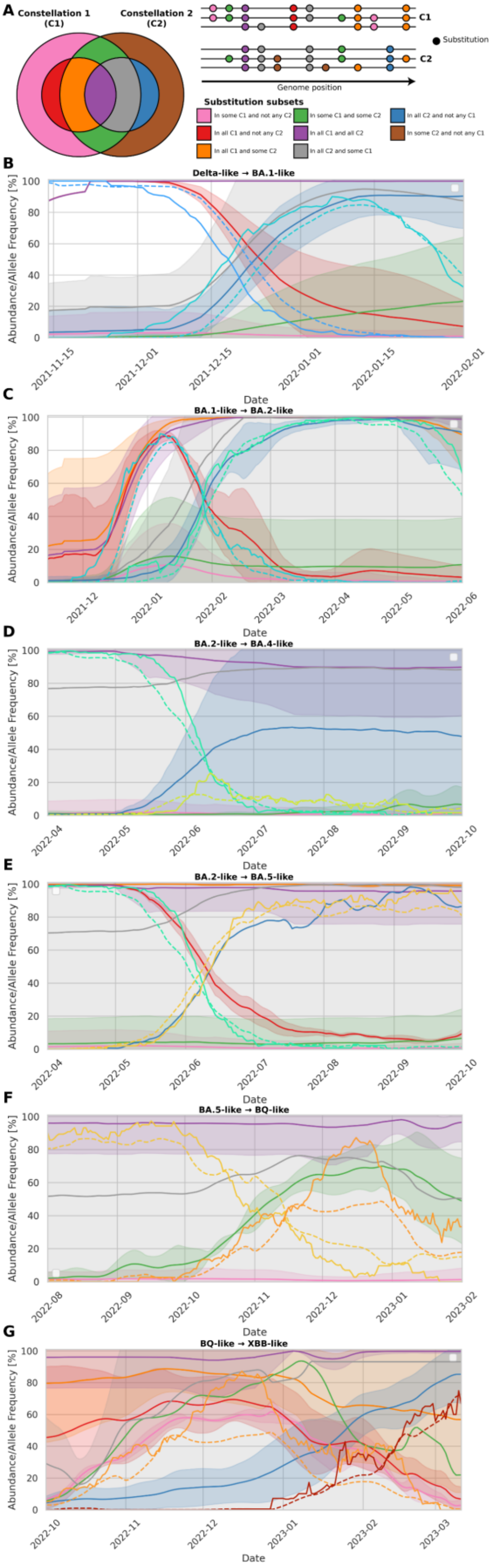

**Figure S5. Set-based substitution tracking of SARS-CoV-2 constellation transitions**

**A**. *Substitution categorisation scheme.* – Left: A Venn diagram depicting how various nucleotide substitutions were uniquely categorized between two constellations (C1 and C2) that participate in a transition. Right: A representation of how these substitutions may appear within multiple lineages of the constellations along the genome, with small circles representing substitutions and are coloured according to how they would be placed into the Venn diagram to the left. The subset colours and categories detailed in the legend are as follows: Pink: Substitutions present within some lineages of the first constellation (C1) but absent in the second constellation (C2). Red: Substitutions found in every lineage in C1 as well as no lineages of C2  Orange: Substitutions found in every lineage of C1 but also present within some lineages of C2. Green: Substitutions found in some lineages in both constellations. Purple: Substitutions found in all lineages defining both constellations.  Grey: Substitutions found in every lineage of C2 but also present within some lineages of C1. Blue: Substitutions found in every lineage in C2 as well as no lineages of C1. Brown: Substitutions present within some lineages of C2 but absent in C1. **B-G**: *Substitution set allele frequency dynamics within the wastewater for the Delta-like→BA.1-like (B), BA.1-like→BA.2-like (C), BA.2-like→BA.4-like (D), BA.2-like→BA.5-like (E), BA.5-like→BQ-like (F), and BQ-like→XBB-like (G) transitions*. The mean allele frequencies for each set are plotted according to their colour scheme with the standard deviation plotted as a semi-opaque shaded area. Note that depending on the particular substitutions belonging to each constellation, certain subsets are empty and are thus not possible to plot (e.g., red in BA.2-like→BA.4-like and blue in BA.5-like→BQ-like).

#### Quantitative assessment of temporal shifts between WW and Pillar 2 sequencing in capturing the rise of SARS-CoV-2 constellations

In examining the first three transition periods (see Figure 2D), characterized by high levels of wastewater (WW) sampling and individual sampling, we observed a temporal shift in the curves, despite displaying similar functional forms (Figure S5A). To quantitatively detect these temporal deviations between the rises of SARS-CoV-2 constellation abundances in the time series, we implemented a systematic horizontal curve shifting algorithm. This algorithm successfully determines the least erroneous shift between each SARS-CoV-2 constellation in these transitions, comparing them to Pillar 2 samples to identify the apparent shift in detection for the rise of the incoming major constellations.

Pillar 2 sequencing, representing broad, community-level sampling, was selected as the reference group due to its significantly higher sampling rates over its period of activity (Figure 2E) and its capacity to mitigate potential biases associated with Pillar 1 sampling within the healthcare system. Unlike Pillar 1 sampling, which often focuses on patients and frontline workers, Pillar 2 sequencing extends testing to the wider population, providing a more representative picture of SARS-CoV-2 variant dynamics across communities in NI.

Our analysis revealed that based on the WW time series derived from Freyja demixing and the Pillar 2 timeseries derived from consensus sequences, there were temporal differences, where wastewater lagged behind Pillar 2 sequencing by 7 days for the rise of BA.1-like (Figure S3B), 3 days for the rise of BA.2-like (Figure S5C), but led by 5 days for the rise of BA.5-like (Figure S5D) constellations.

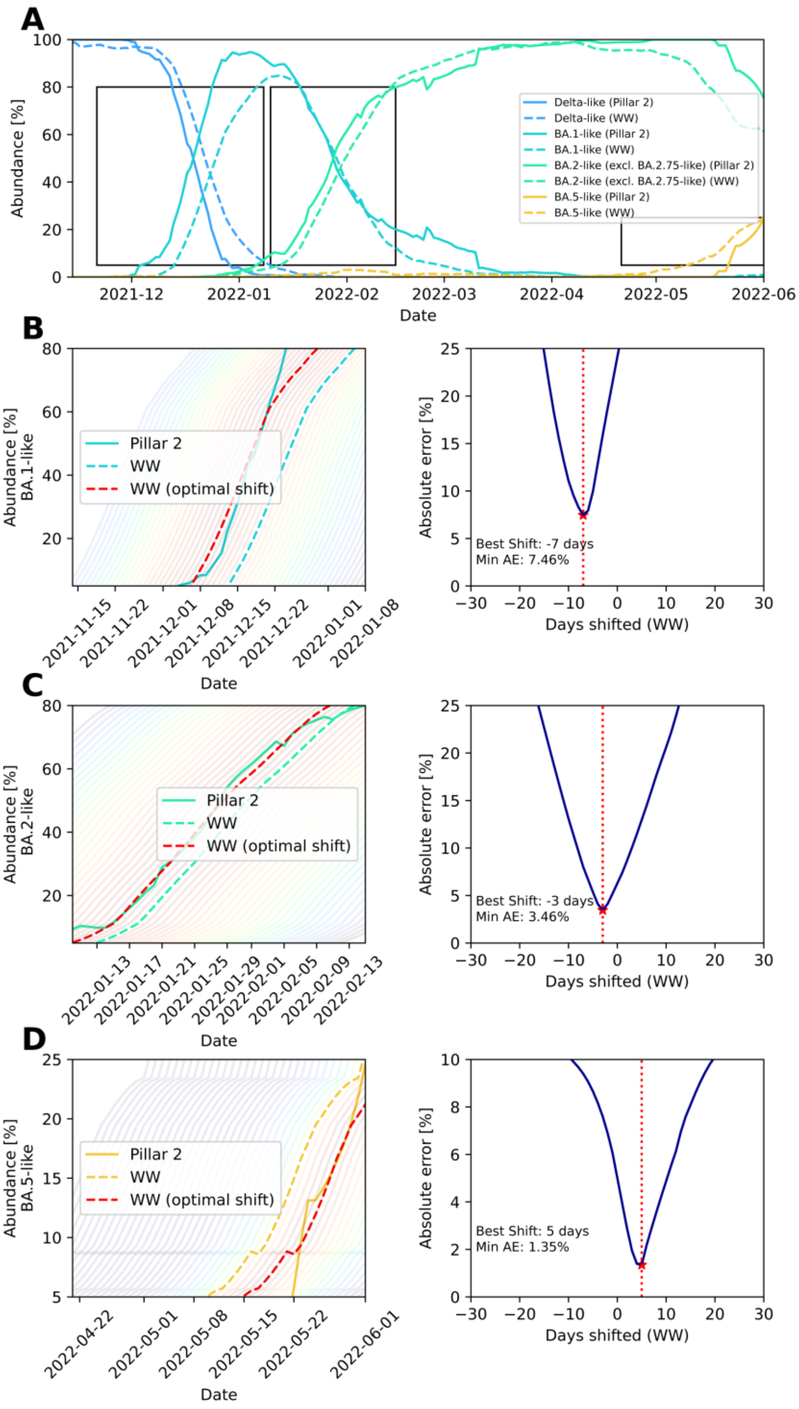

**Figure S6. Temporal deviations and optimal alignment analysis between WW and Pillar 2 (individual) sequencing**

A: *The first 3 transition periods between major SARS-CoV-2 constellations during the WW and Pillar 2 sequencing programmes*. – We subsetted the major constellation groups from the constellation time series depicted in Figure 2A, focusing on the overlap of the Pillar 2 and WW sequencing programmes. Abundances from the individual sequencing programmes are represented by solid lines, whereas those determined from the wastewater (WW) sequencing programme are depicted as dashed lines. Black rectangles denote the emphasized temporal ranges in the lower panels (B-D). B-D: *Temporal shifting scans to determine shift times that maximise agreement between Pillar 2 individual sampling and WW for the rise of the BA.1-like (B), BA.2-like (C), and BA.5-like (D) SARS-CoV-2 constellations.* – The left panels display the optimal alignment algorithm for the rise of each SARS-CoV-2 constellation, where the semi-transparent rainbow colour scale depicts the goodness of fit (indigo = worst-fitting of the scan, red = optimal fit). The right panels show the optimisation metric, with the absolute errors obtained for each shift value depicted in blue. The optimal temporal shift is annotated and highlighted by a vertical red dotted line.

#### Substitution sets tracked in Figure S5

Regarding the substitution sets in our analyses, the set of substitutions will depend on the two sets being compared see Figure S5A. Thus, the following table can be used, keeping in mind the transition occurring. The empty set is depicted as {}. Each transition is depicted as C1→C2 in Figure S5.

**Table S6. Substitution sets tracked in transitions (see Figure S5).**

| Constellation 1 (C1) | Constellation 2 (C2) | All C1 not C2 (red) | All C2 not C1 (blue) | Some C1 not C2 (pink) | Some C2 not C1 (brown) | | All C1 some C2 (orange) | All C2 some C1 (gray) | All C1 and all C2 (purple) | Some C1 and some C2 (green) |
|---|---|---|---|---|---|---|---|---|---|---|
| Delta-like | BA.1-like | G210T C21618G T22917G C23604G C25469T C27752T A28461G G28881T G29402T G29742T | C10449A C21762T C21846T G22578A T22673C C22674T T22679C C22686T G22992A A23040G A23055G T23075C C23525T T23599G C23604A C23854A C24130A A24424T T24469A C25000T C25584T A27259C A28271T C28311T G28881A G28882A G28883C | [illegible] | [illegible] | | {} | C10029T C15240T A23063T G23948T C27807T | C241T C3037T C14408T C22995A A23403G | C5730T C11455T C12513T G22599A G22813T A23013C C23934T G27762T G28378T |
| BA.1-like | BA.2-like | C15240T C21762T C21846T T22673C C24130A | G4184A C4321T C9344T A9424G C9534T C10198T G10447A C12880T C15714T C17410T C19955T A20055G C21618T T22200G A22688G G22775A A22786C C26060T C26858T G27382C T27384C A29510C | [illegible] | [illegible] | | C22674T C22995A A23040G C23604A | A18163G G22813T T22882G C26270T C26577G G26709A | C241T C3037T C10029T C10449A C14408T G22578A T22679C C22686T G22992A A23055G A23063T T23075C A23403G C23525T T23599G C23854A G23948T A24424T T24469A C25000T C25584T A27259C C27807T A28271T C28311T G28881A G28882A G28883C | C3602T C4582T C5183T G8393A G19684T G22580A G22599C G22898A A23013C C23673T C26078T C28472T |
| BA.2-like | BA.4-like | {} | T23018G C28724T | [illegible] | [illegible] | | {} | T670G C2790T G12160A C22674T T22917G C22995A A23013C C23604A A27383T | C241T C3037T G4184A C4321T C9344T A9424G C9534T C10029T C10198T G10447A C10449A C12880T C14408T C15714T C17410T A18163G C19955T A20055G C21618T T22200G G22578A T22679C C22686T A22688G G22775A A22786C G22813T T22882G G22992A A23055G A23063T T23075C A23403G C23525T T23599G C23854A G23948T A24424T T24469A C25000T C25584T C26060T C26270T C26577G G26709A C26858T A27259C G27382C T27384C C27807T A28271T C28311T G28881A G28882A G28883C A29510C | C13329T A22001G G22017T G22599C G22894C T22942G C28435T |
| BA.2-like | BA.5-like | C26858T A27259C G27382C T27384C | G26529A | [illegible] | [illegible] | | C14408T T22200G A28271T | T670G C2790T G12160A C22674T T22917G A23013C C23604A | [illegible] | [illegible] |
| BA.5-like | BQ-like | {} | {} | [illegible] | {} | | {} | C1931A T2954C C11750T C14408T G16935A T22200G C22995A T23018G C27889T A28271T C28312T G28681T | [illegible] | T14257C A17039G G22599C A22893C T22942A |
| BQ-like | XBB-like | C1931A T2954C G12160A G16935A T22200G G26529A G28681T | A405G C9866T G15451A C15738T T15939C T17859C A19326G T21810C C22000A C22109G T22200A G22577C C22664A G22895C T22896C G22898A T22942G T23019C T23031C C25416T A26275G C26858T A27259C G27382C A27383T T27384C | T14257C A17039G T22942A | [illegible] | | C11750T T22917G C22995A T23018G T23075C C23604A G23948T C27889T A28271T C28312T | G22599C | [illegible] | A22893C |

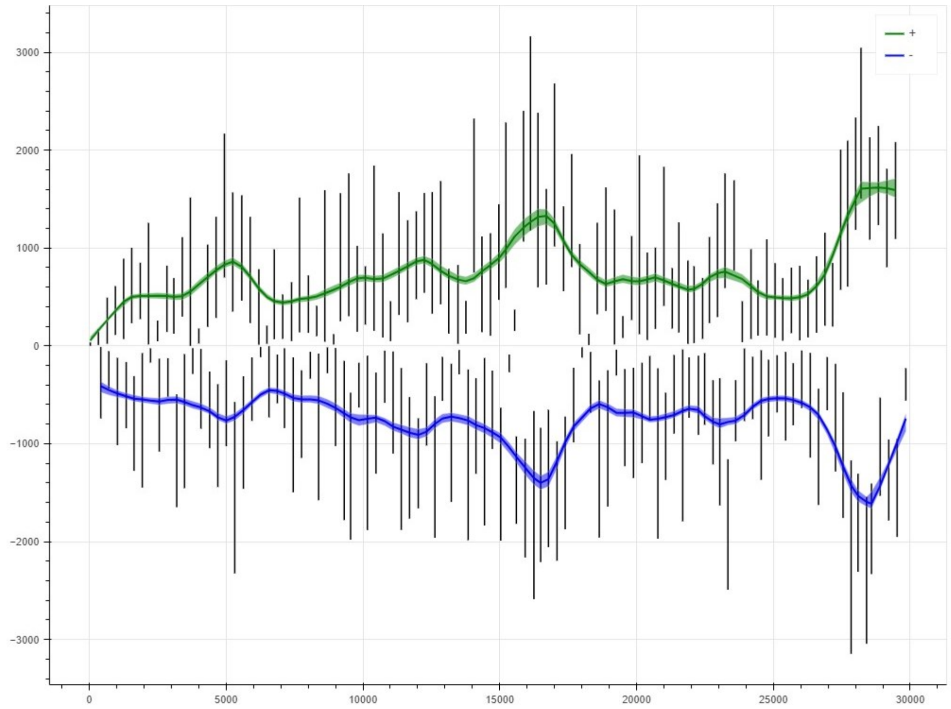

**Figure S7. Coverage of ARTIC V4 amplicon primers for exemplary WW WGS.**

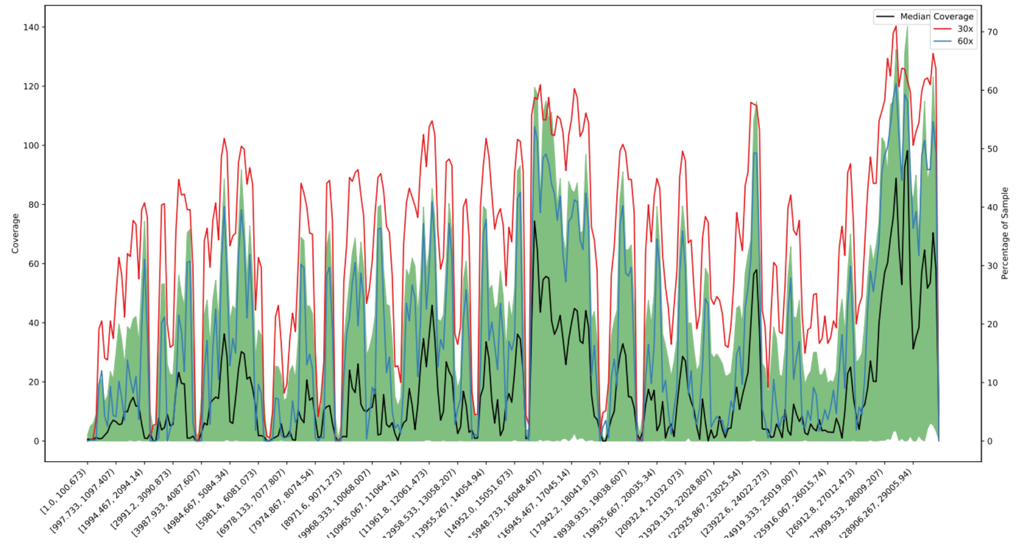

**Figure S8. Genome coverage for an exemplary subset of the WW WGS.**
